# Recent and forecast post-COVID trends in hospital activity in England amongst 0 to 24 year olds: analyses using routine hospital administrative data

**DOI:** 10.1101/2021.02.11.21251584

**Authors:** Joseph Ward, Dougal Hargreaves, Marie Rogers, Alison Firth, Steve Turner, Russell Viner, On behalf of the Royal College of Paediatrics and Child Health Paediatrics 2040 Data Working Group

**Affiliations:** UCL Great Ormond St. Institute of Child Health, London; Department of Primary Care and Public Health, Imperial College London; Royal College of Paediatrics and Child Health, London; Child Health, University of Aberdeen, Aberdeen and the Royal Aberdeen Children’s Hospital, NHS Grampian, Aberdeen

## Abstract

**Background:** Increasing hospital use in the past decade has placed considerable strain on children and young people’s (CYP) health services in England. Greater integration of healthcare may reduce these increases. We projected CYP healthcare activity out to 2040 and examined the potential impact of integrated care systems on projected activity.

**Methods:** We used routine administrative data (Hospital Episode Statistics (HES)) on emergency department (ED) attendances, emergency admissions and outpatient (OP) attendances for England by age-group for 0-24 year olds from 2007 to 2017. Bayesian projections of future activity used projected population and ethnicity and future child poverty rates. Cause data were used to identify ambulatory-care-sensitive-conditions (ACSC).

**Findings:** ED attendances, emergency admissions and OP attendances increased in all age groups from 2007 to 2017. ED and OP attendances increased 60-80% amongst children under 10 years. ACSC and neonatal causes drove the majority of increases in emergency admissions. Activity was projected to increase by 2040 by 50-145% for ED attendances, 20-125% for OP attendances and 4-58% for total admissions. Scenarios of increasing or decreasing child poverty resulted in small changes to forecast activity. Scenarios in which 50% of ACSC were seen outside hospital in integrated care reduced estimated activity in 2040 by 21.2-25.9% for admissions and 23.5-30.1% for ED attendances across poverty scenarios amongst infants.

**Interpretation:** The rapid increases in CYP healthcare activity seen in the past decade may continue for the next decade given projected changes in population and child poverty, unless some of the drivers of increased activity are addressed. Contrary to these pessimistic scenarios, our findings suggest that development of integrated care for CYP at scale in England has the potential to dramatically reduce or even reverse these forecast increases

**Funding:** Nil funding obtained.

**Research in context:** *Evidence before this study:* There has been marked increases in hospital use (inpatient, outpatient and emergency department (ED)) by children and young people (CYP). Search of the PubMed database using the search terms: (((((“child”[MeSH Major Topic]) OR (“adolescent”[MeSH Major Topic])) OR (“infant”[MeSH Major Topic]))) AND ((healthcare use[Text Word])) OR (emergency admission[Text Word])) AND (united kingdom[Text Word]). Drivers of increased activity include population growth and sociodemographic factors, help-seeking behaviour, growth in medical knowledge and capability, and by factors within the health system. Additional factors in child health include increased survival of premature neonates and those with congenital conditions and rising parental expectations of modern medicine. Previous studies have shown that ambulatory-care-sensitive-conditions (ACSC) are responsible for much of the increase in CYP emergency activity in England and Scotland.

*Added value of this study:* This is the first study to use existing data to project possible future scenarios for CYP healthcare activity out to 2030 and 2040 in any country. Our future scenarios are based upon authoritative projections for population, ethnic diversity and child poverty in England and allow us to estimate the potential impact of integrated care scenarios in which ACSC are treated outside hospital. We show that future projected CYP activity is very high if mitigations such as integrated care are not instituted in England.

*Implications of all the available evidence:* Healthcare activity has grown dramatically over the last decade in CYP, largely due to ACSC and the consequences of premature delivery. Projections to 2040 suggest that similar increases are likely over the next 2 decades without action to reduce child poverty and implementation of integrated care at scale in the NHS.

## Background

There has been marked increases in hospital use by children and young people (CYP) in England over the past decade.^1-3^ Healthcare activity is driven by health need, population growth and sociodemographic factors, by patient expectations and help-seeking behaviour, by the growth in medical knowledge and capability, and by factors within the health system including increased fragmentation.^4^ Specific paediatric factors increased survival of premature neonates and those with congenital conditions leading to an increase in disability and complexity for paediatric services, broadening perspectives on the role of medicine in developmental and behavioural issues, allied with rising parental expectations of modern medicine. Sociodemographic change may also contribute including increasing deprivation in the CYP population.^5^ Limited capacity in primary care and lack of anticipatory preventive have also been identified as drivers of increased CYP emergency activity care in England.^1,6,7^ whilst lower quality of secondary care has been linked to higher emergency admission rates for long-term conditions.^8^

Efforts to reduce the apparently inexorable rises in healthcare activity, particularly emergency activity, have focused recently on greater integration of care across different layers of healthcare and across health, social care and education sectors.^9,10^ Integrated care aims to provide more services outside hospitals at the interface between primary and secondary care and increase integration across health and social care, with increased attention to prevention. In adults, better integrated care improves patient satisfaction, perceived quality of care, and improves access to services.^10^ Improved integration of care for CYP across is a key element of plans to improve outcomes for CYP in the UK.^11-13^ A major component of better integrated care is reducing unnecessary hospital attendances and providing better quality care closer to home when possible^9,11,14^ Although the impact of increased integration across primary and secondary services on healthcare activity for CYP has achieved relatively little attention, there is evidence from small-scale studies this can reduce both emergency and planned activity.^1,15^

The Royal College of Paediatrics and Child Health (RCPCH) launched the Paediatrics 2040 Project in 2019 to inform long-range planning of healthcare and need for CYP. The aim of this paper is to examine CYP healthcare activity over the past decade and use this to forecast trends to 2030 and 2040, given potential changes in child poverty and increased health system integration. We hypothesized that the most visible outcome of increased integration would be reduction of emergency hospital activity due to management of ambulatory-care sensitive conditions (ACSC) in out-of-hospital settings. The COVID-19 pandemic provided a real-world case study of sudden rapid shift in some of the drivers of healthcare activity leading to marked reductions in healthcare activity^16^ together with rapid innovation in the delivery of care.^17^ COVID-19 presents health-care systems with opportunities to preserve and build upon changes made during the pandemic.

## Methods

### Data sources

Routine administrative data (Hospital Episode Statistics (HES) data provided by NHS Digital)^18^ were obtained by age-group for 8 days to 24 year old for inpatient admissions, outpatient (OP) attendances and emergency department (ED) attendances in England from 2007 – 2017. Attendance rates were calculated using mid-year population denominators provided by the Office for National Statistics (www.ons.gov.uk). Attendances were categorised by region, deprivation (Index of Multiple Deprivation (IMD) quintile) and type: emergency admission or planned admission (Elective or Day case admissions) or Transfer admissions. For these analyses we did not include the very small numbers of transfer admissions as these were a mix of emergency and elective admissions transferred between hospitals. Cause data were only available for inpatient admissions; International Classification of Diseases 10 (ICD10) codes were mapped to level 3 causes of the Global Burden of Disease 2017 cause hierarchy^19^ and regrouped to create 48 cause-clusters more clinically relevant for CYP (Appendix Table 1). Emergency presentations manageable in integrated care systems outside the hospitals (Ambulatory Care Sensitive Conditions (ACSC)) were identified using ICD-10 codes provided by Cecil et al.^7^ and NHS Digital^20^ including upper respiratory tract infections (URTI), lower respiratory infections, diarrhoea, asthma/wheeze, ENT conditions, skin conditions, epilepsy and common infections (e.g. vaccine preventable) and presentations for non-specific viral infections, non-specific symptoms and somatic complaints (e.g. headache). Note that OP attendances data before 2008 and ED attendances data before 2012 are considered provisional as coverage was not fully national.

### Analyses

Projections to 2040 used Bayesian probabilistic models to forecast outcome values simulated from the posterior predictive distribution. Projections used the *Bayesmh* commands and *Bayespredict* commands in Stata 16 (StataCorp, College Station TX), using weakly informative priors. Three chains were run for each model, each consisting of a Markov chain Monte Carlo (MCMC) sample size of 30,000. Separate models were run for each age group with 99% credible intervals (CI) forecast around each mean forecast. Models were restricted to all-cause activity for ED attendances, emergency admissions and OP attendances to minimise modelling. Planned admissions (elective and day-case admissions) were not modelled as they did not change across the last decade. We therefore estimated planned admission rates from 2018 to 2040 as being unchanged from the mean of 2015-2017. Total admission rates from 2018 to 2040 were then estimated by addition of emergency and planned admission rates.

Models included a limited set of explanatory variables known to be associated with activity in CYP for which historical data and authoritative future forecasts are available, including population and ethnicity^21^ and proportions of children in relative poverty (<60% of median income, after housing costs).^22^ We used short-term forecasts of a rise in child poverty from 29.5% in 2017 to 36.5% by 2021^5^ to examine three future poverty scenarios including i) ‘stable poverty’, in which child poverty continues unchanged from 2017 to 2040 at 29.5%; ii) ‘decreasing poverty’, in which child poverty falls to 20% by 2021 and remain at this level to 2040; and iii) ‘increasing poverty’, in which child poverty rises to 36.5% by 2021 and remains at this high level to 2040.

We also estimated future scenarios in which health system reorganisation towards integrated care after the COVID-19 pandemic resulted in reductions in CYP ED attendances and emergency admissions due to ACSC being managed outside of hospitals. These scenarios began with the observed reduced activity during the pandemic from April to August 2020 in England (25% in emergency admissions and 50% in ED attendances and elective admissions; personal communication, Simon Kenny, NHS England). These scenarios were estimated by calculating the impact of hypothesised reductions in activity due to ACSC on the stable poverty forecasts.

First, the ‘COVID shock’ scenario hypothesised there was no significant reorganisation of healthcare and that activity returned to predicted levels in a linear fashion over 2020-2023. Second, two integrated care scenarios hypothesize that the reductions during the pandemic are maintained as health service change is implemented to maintain innovation and implement integrated care at scale. The ‘Fully integrated care’ scenario assumed that 100% of ACSC were managed outside the hospital system, and the ‘Moderate integrated care’ scenario assumed 50%. To calculate these we identified the proportion of emergency admissions that were ACSC for each age-group in 2017 and assumed this proportion remained constant to 2040. The ‘increasing poverty’ scenario obviated the need for a separate scenario for increased poverty and disruption in the social determinants of health post-COVID. We did not estimate COVID impacts upon OPD attendances due to lack of information on drivers of increased activity and impacts of COVID-19.

### Ethics

Analyses were approved by the London-Brent NHS Research Ethics Committee (ref: 18/LO/1267).

## Results

Trends in ED attendances and admissions from 2007-2017 are shown by age-group in Figure 1 and Appendix Table 2). ED attendances and emergency admissions increased in all age groups, most dramatically in under ‘10s. There were small increases in Day-Case admissions in all age-groups with concomitant falls in elective admissions, with overall planned admissions stable.

**Figure 1:**
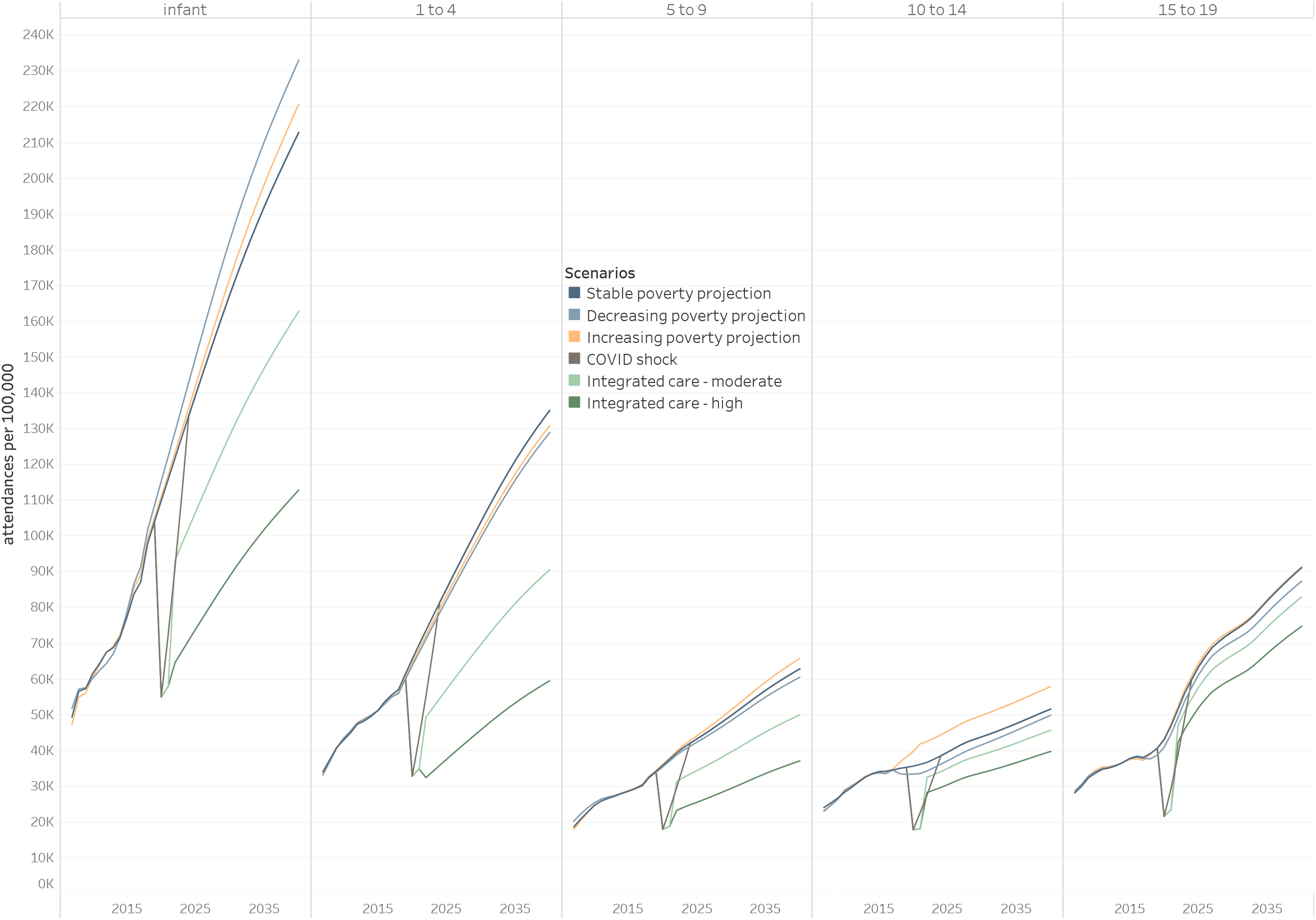
Trends in emergency attendances and emergency and elective admission by age in England, 2007-2017 (both sexes)

Regional trends in ED attendances and emergency admissions were similar to the England average, although with marked disparities between regions that changed little over the decade (Figure 2 and Appendix Figure 1). ED attendances were consistently higher than the England average across the period in London, the North-East and the North-West of England and lower in all other regions. Similarly for emergency admissions, the North-East, North-West and West-Midlands had rates consistently higher than for England however London had the lowest rate in each age group.

**Figure 2.**
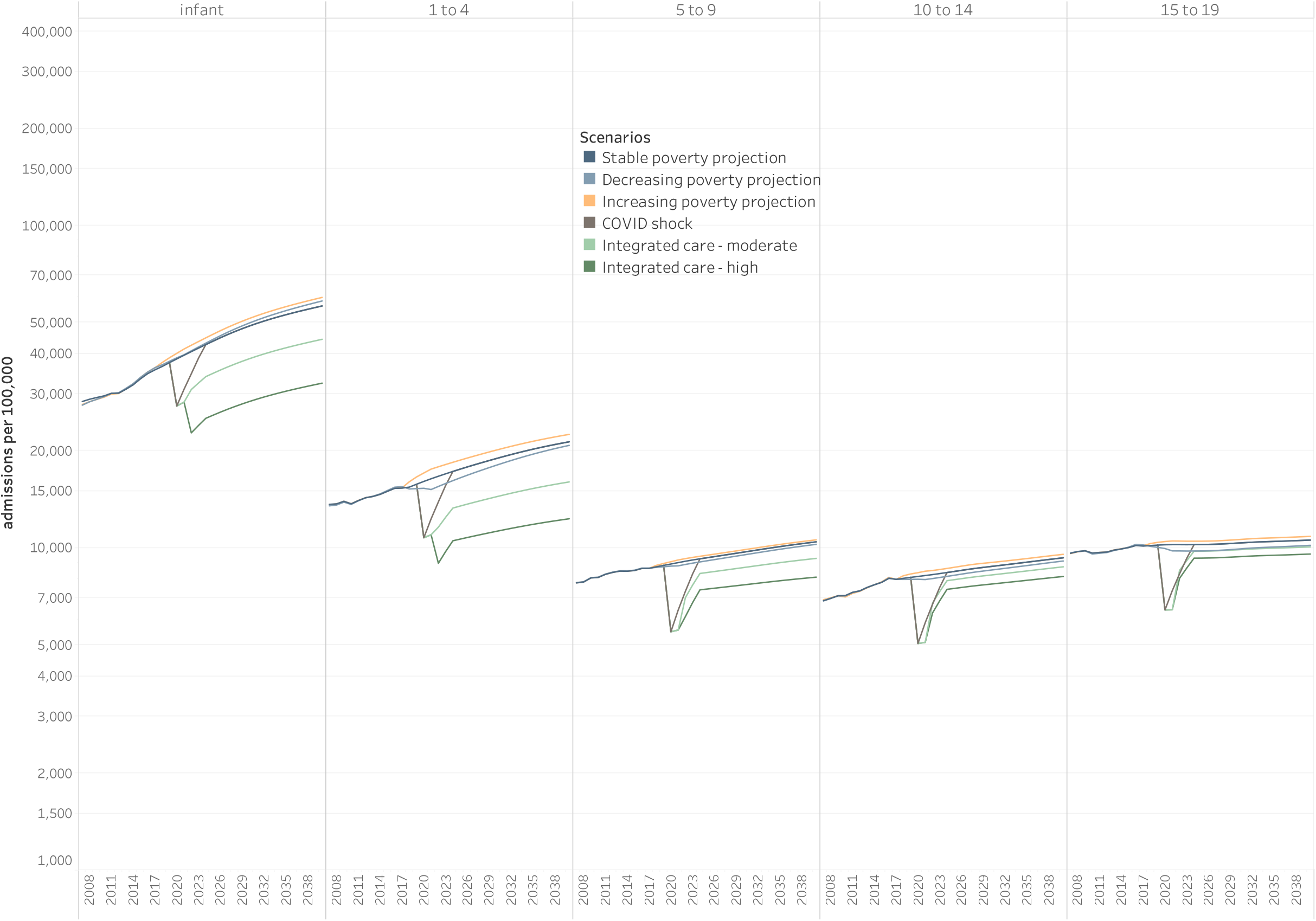
ED attendances and Emergency Admission rates by English region for 0-4 year olds, 2007-2017 (both sexes)

There were strong social gradients in activity across age-groups, with CYP from the most deprived groups being most likely to attend ED and be admitted (emergency or elective) (Appendix Figure 2) across the study period.

Causes of emergency admissions amongst 0-4 year olds are shown in Figure 3, categorised as ACSC or other causes. Proportional changes by cause are shown in Appendix Table 3. Eight of the top ten causes in 2017 were ACSC. The top five causes (the consequences of prematurity together with four ACSC (upper and lower respiratory infections, other common infections and non-specific fevers) were also those which increased most across the decade, with little change in the majority of other causes.

**Figure 3.**
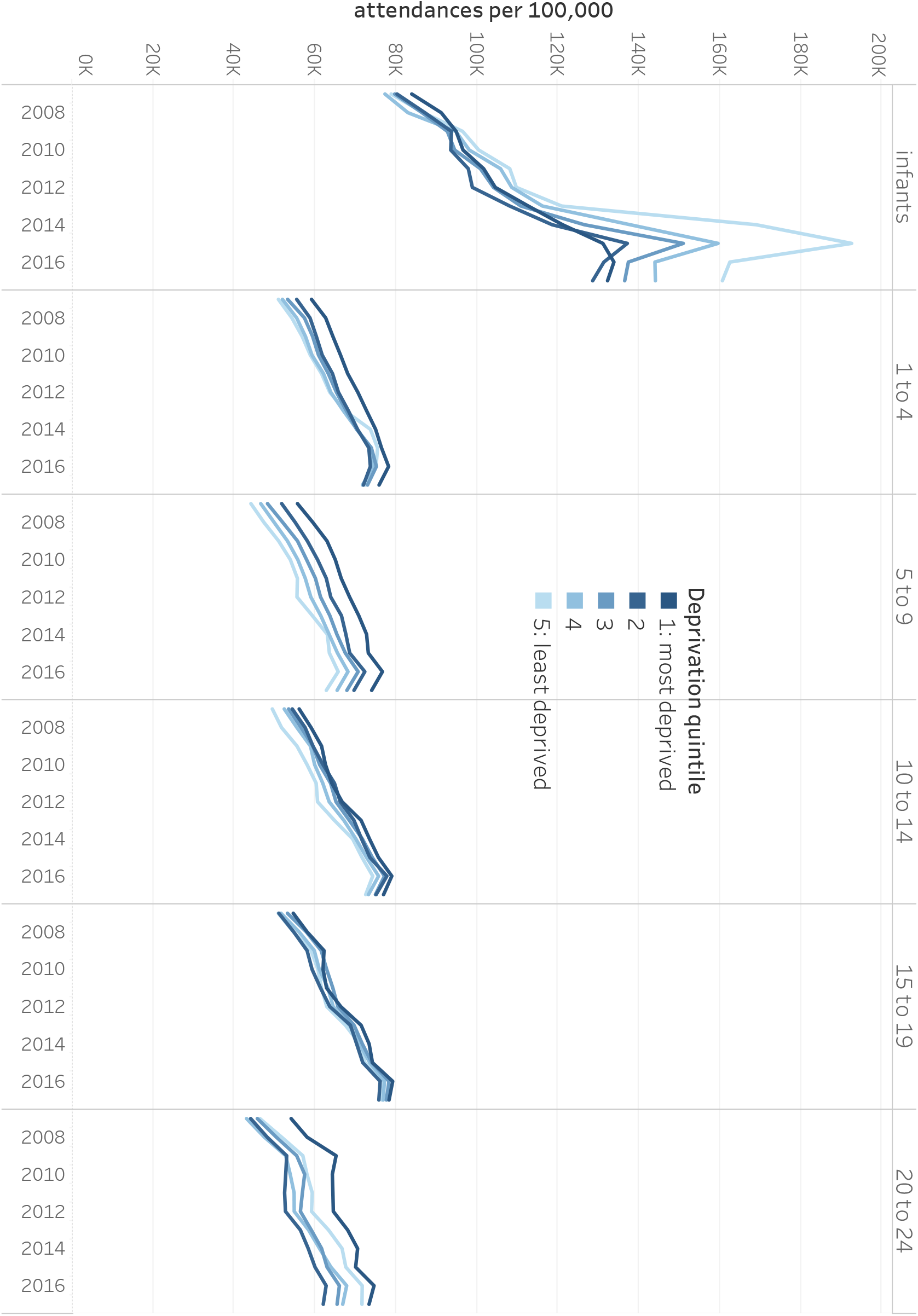
Trends in emergency admissions by cause amongst 0-4 year olds in England, 2007-2017 (both sexes) Note: Only causes responsible for ≥2% of total burden for each year and age-group are shown to aid clarity.

Main causes of emergency admissions amongst 5-24 year olds are shown in Appendix Figures 3 and 4. Injuries were the commonest cause in those over 5 years but diminished across the decade in each age-group. Amongst 5-9 year olds, ACSC formed the majority of the other main causes, with notable increases in infectious cause admissions, particularly fever/nonspecific viral infections, over the decade. In 10-24 year olds, the main causes were nonspecific symptoms, neurodevelopmental causes, gastrointestinal and renal disorders, with neurodevelopmental conditions the only cause to increase notably across the study period. Increases in infectious causes were also visible in 10-24 year olds however they made up only a small proportion of admissions.

**Figure 4.**
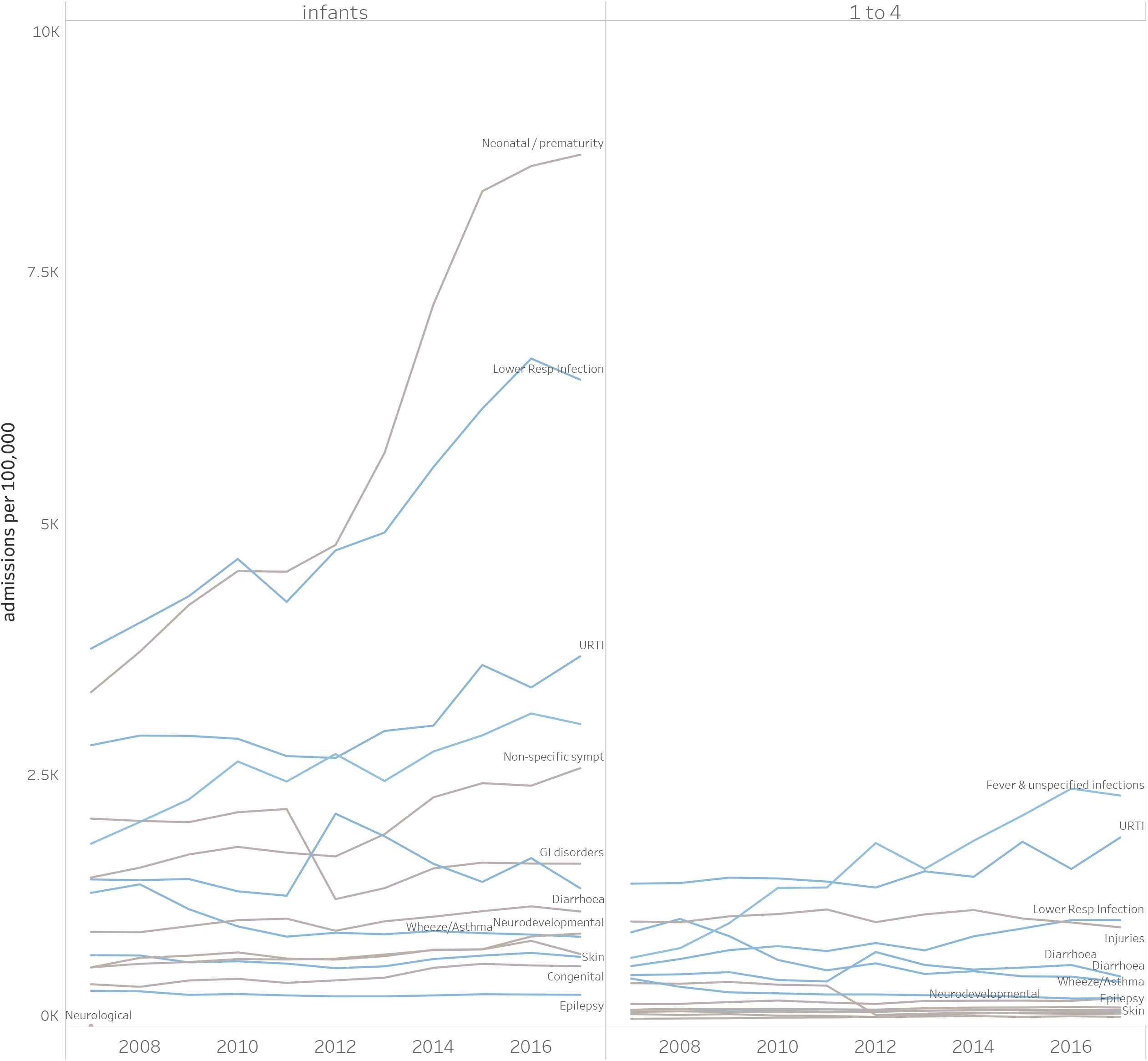
Outpatient attendances by age and deprivation quintile in England, 2007 to 2017

In 2017, ACSC made up 47% of emergency admissions amongst infants, 66% amongst 1-4 year olds, 41% of 5-9 year olds and 17-23% of adolescents (Appendix Figure 5). The proportion of infant emergency admissions that were ACSC was lowest in London (40%) but otherwise varied little by region (Appendix Figure 6).

**Figure 5.**
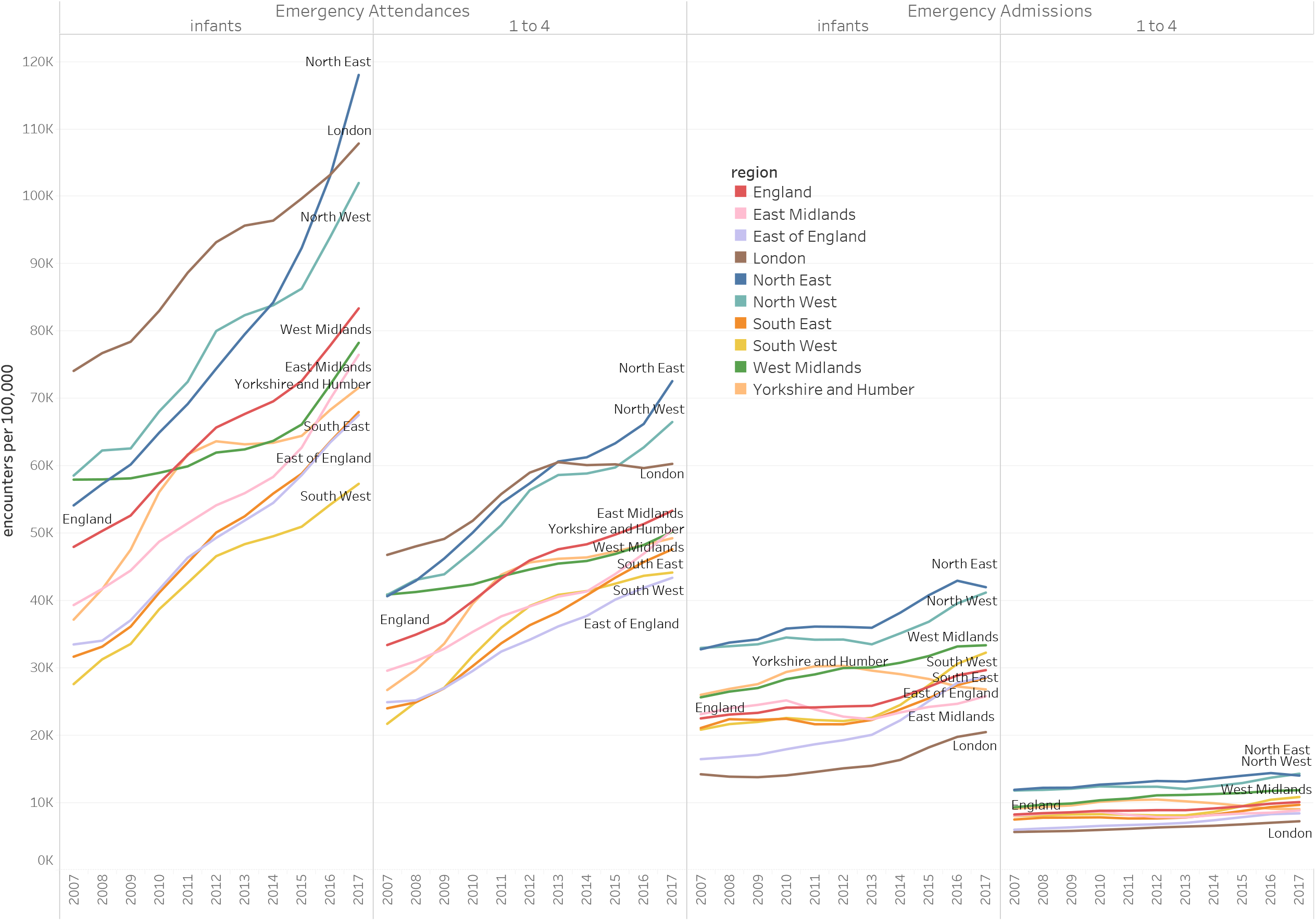
Projections of total admission rates in England to 2040 according to poverty and integrated-care scenarios, by age-group

Elective admissions from most causes declined across the decade in all age-groups (Appendix Figures 7 8). Admissions in 0-4 year olds were dominated by congenital, neonatal, gastrointestinal and chronic respiratory causes, whilst injuries and a range of other non-communicable causes dominated in those 5-24 years.

The largest increase in OP attendances rates was amongst infants (72% from 2007-2017) whilst increases in all other age-groups were 34-47% (Appendix Table 4). These changes varied by deprivation; amongst infants the social gradient reversed between 2007 and 2017, with attendances increasing most in the least deprived quintile (144% compared with 58% in most deprived; Figure 4). A similar pattern of greatest increases amongst the least deprived was seen in 20-24 year olds, whilst attendances were highest amongst the most deprived quintile in all other age-groups.

Projections of future CYP healthcare activity from 2018 to 2040 in scenarios of stable, increasing and decreasing poverty are shown in Figure 5 for total admissions and Figure 6 for ED attendances (also see Appendix Tables 5 and 6 and Appendix Figures 9 to 11).

**Figure 6.**
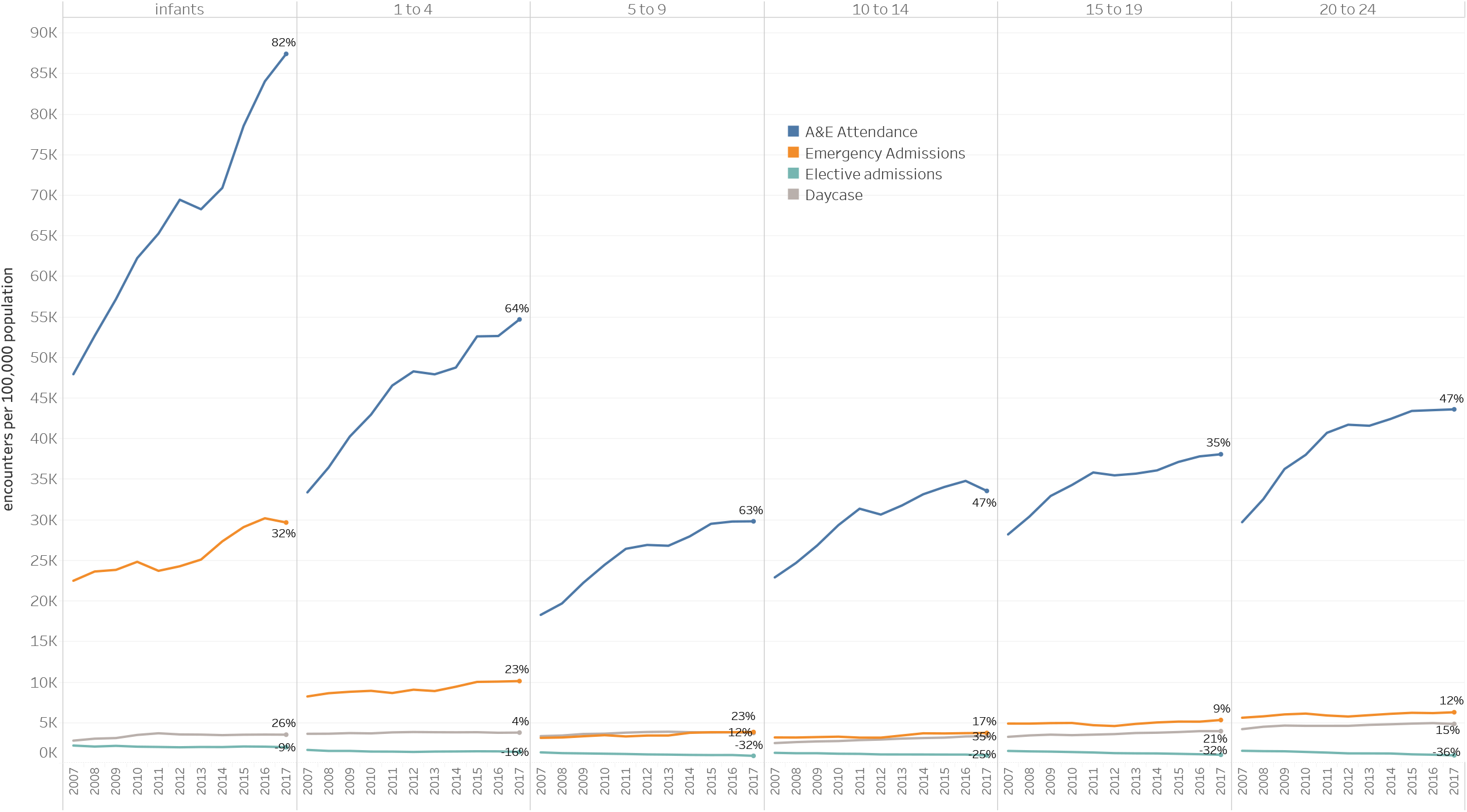
Projections of ED attendances rates in England to 2040 according to poverty and integrated-care scenarios, by age-group

In the stable poverty scenario, total admissions were forecast to increase by 2040 by 58% in infants, 39% in 1-4, 21% in 5-9, 17% in 10-14 and 4% in 15-19 year olds, with ED attendances similarly increasing by 144%, 143%, 107%, 49% and 140% respectively. In the decreasing poverty scenarios, forecast admission rates were lower by 2 to 5% than for stable poverty in total admissions and ED attendances in all age groups except amongst infants, where 99%CI overlapped markedly with stable poverty scenarios. Similarly in the increasing poverty scenarios, total admission rates and ED attendances were 3-5.5% higher than in the stable poverty scenarios except in infants where 99% CI overlapped markedly.

Post-COVID scenarios are also shown in Figures 5 and 6, with reductions compared with the poverty scenarios shown in Appendix Table 7. The temporary COVID shock scenario had no impact upon forecast long-term activity. The impact of moderate or high levels of health system integration post-COVID appears highest amongst those under 10 years. Amongst infants, the moderate integration scenario reduced estimated activity in 2040 by 21.2-25.9% for admissions and 23.5-30.1% for ED attendances across poverty scenarios, whilst the high integration scenario decreased activity 42.4-45.8% and 47.0-51.6% respectively. Reductions of a similar order were seen for 1-9 year olds, whilst reductions amongst 10-19 year olds were considerably smaller.

OP attendances projections to 2040 in different poverty scenarios are shown in Appendix Figure 12 and Appendix Tables 8-10. In the stable poverty scenario, OP attendances rates were projected to increase to 2040 by 89% compared with 2016 in infants, 21% in 1-4 year olds, 90% in 5-9 year olds, 57% in 10-14 year olds and 124% in 15-19 year olds. There was very notable overlap of 99% CI for decreasing and increasing poverty scenarios with the stable poverty scenario in all age-groups aside from 1-4 year olds. Amongst 1-4 year olds, increasing poverty led to a 32% increase whilst decreasing poverty led to only a 6% increase on 2016.

## Discussion

Trends in hospital activity in 0-24 year olds over the past decade show marked rises in emergency activity (admissions and ED attendances) and OP attendances, particularly amongst under ‘10s but visible across all age-groups. Increases were greatest for ED and OP attendances, both of which increased over 70% amongst infants. These patterns were visible across all regions, although increases in the North-East and North-West were consistently above the England average for both emergency admissions and ED attendances, with East of England, South-West and South-East consistently below England average. London uniquely showed high ED attendances but low emergency admissions, suggestive of lower translation of attendance into admission. Trends appeared largely uniform across levels of deprivation, with the exception of OP attendances in infants, where the social gradient reversed over the decade due to dramatic rises in the amongst the least-deprived. The reasons for this are unclear but may reflect changes in parental help-seeking behaviour. Cause-level analysis for emergency admissions suggested that the changes in emergency activity over the past decade seen particularly in young children have been driven predominantly by rises in a small number of conditions, particularly non-specific fevers, viral infections and respiratory infections. Each of these are ACSC that are potentially manageable outside hospital settings. It is unclear whether the rise in these presentations are due to a greater awareness of the dangers of sepsis, rises in parental anxiety, reduced access to primary care or changing thresholds for admission.

The large increases in emergency activity and OP attendances documented across the past decade in young children, of the order of 60-80%, have placed great strain on children and young people’s services in England, as the children’s workforce and service structure has not increased to match it. Workforce data from the RCPCH shows that whilst consultant paediatrician numbers in England increased 47% between 2007-2017, most of the increase was in specialist rather than general or acute paediatricians, whilst proportions of consultants working less-than-full-time increased.^23^ The RCPCH estimated in 2019 that an additional 642 WTE consultant paediatricians were needed to meet demand in England.^24^ At the same time that emergency admissions were rising, numbers of hospital beds for children in England fell 16.4% from 5315 in 2007/08 to 4441 in 2016/17,^25^ further illustrating increasing strain on the system.

Our most conservative scenario, in which there are no substantial changes in terms of child poverty or health system organisation over the next two decades, predicts increases of 50-145% in ED attendances and 20-125% increases in OP attendances. An alternative scenario where policy action reduces child poverty significantly over the next two decades has a beneficial impact upon these forecasts, although the forecast impact is less than 5% for all activity. These changes will require significant additional workforce and health services resources.

In contrast, integrated care scenarios dramatically reduced projected future activity across all admission types, suggesting markedly lower needs for future additional workforce and capacity. Managing all ACSC outside the hospital system (high integrated care scenario) could potentially reduce total admissions to at or below 2007 levels by 2040; reductions in increases in ED attendances are predicted to be less dramatic, achieving levels close to those of 2017 over the next two decades. Whilst redirection of 100% of ACSC is unlikely to be easily achieved, the more pragmatic scenario of managing 50% of ACSC outside hospitals (moderate scenario) offers still valuable reductions in forecast activity. Forecast benefits are greater amongst younger children due to higher proportions presenting with ACSC.

We did not model the impact of increased integration of care on OPD attendances. However small-scale studies suggest that up to 40% of OP attendances by CYP might be avoided by more integrated care.^15^

Our findings that viral infections and particularly respiratory infections in young children were the main causes of rapid increases in emergency activity over the past decade are similar to findings from studies from Scotland^26^ and England.^27^ The short-term impact of COVID-19 modelled here is consistent with impacts of the pandemic on emergency activity in Scotland^16^ and Germany.^28^ We are not aware of any other published studies that have attempted to forecast future CYP healthcare activity in England or internationally. Large-scale forecasting efforts, such as those produced by the Institute of Health Metrics and Evaluation (IHME) for the Chief Medical Officer’s 2018 Annual Report,^29^ have been confined to mortality and years of life lost (YLLs) and have not forecast healthcare activity.

### Strengths and limitations

We used a decade of high quality routine hospital administrative data for England to examine trends in total and cause-specific activity for 0-24 year olds. We modified the GBD cause hierarchy to identify cause groups most relevant to children. Our use of Bayesian probabilistic projections, based upon authoritative population forecasts and potential child poverty scenarios, provided a probability distribution of the quantity of interest.

Our findings are subject to a number of limitations. Data for OP attendances and ED attendances are of lower reliability before 2008 and 2012 respectively. Our forecasts are merely possible scenarios for future activity and are necessarily speculative. We were only able to include a very limited number of inputs, which whilst causally related to activity, were themselves forecasts. Our estimates of the impact of greater integration of care were limited to reducing ACSC activity in hospitals and we were not able to examine impacts upon other elements of hospital activity nor on activity in non-hospital settings. It is possible that integrated care models may simply move attendances from hospitals to non-hospital settings. Additionally, these estimates were based upon the assumption that the proportion of ED attendances due to ACSC was similar to the proportion of emergency admissions caused by ACSC. This is likely an under-estimate, given that a greater proportion of less severe cases present to ED than are admitted to hospital. As in any study dependent on ICD-10 coding of causes, misclassification of admission causes may have biased findings although the direction of biases is unclear. In the interests of parsimony, we only ran models for both sexes combined although we recognise that there are substantial differences in admission rates by gender in older adolescents.

### Policy implications

High quality and accessible services for CYP are essential to improving the health of CYP in the UK, a particular need given that health outcomes for CYP rank poorly compared with other wealthy countries.^3^ Health services for CYP are also an important investment in the future development of a country, as poor quality care and unmet healthcare need in childhood is associated with long-lasting effects on mortality, physical and mental health, and healthcare seeking behaviour in adulthood.^8^ Long-term planning is required to enable us to plan and resource such services. Our forecasts suggest that, if drivers of increased activity are not addressed, there will be further rapid increases in CYP emergency and outpatient activity over the next 20 years, requiring significant additional investment in both services and workforce if quality is not to fall. Without concerted action to reduce child poverty, healthcare activity will increase and outcomes worsen. Contrary to these pessimistic scenarios, our findings suggest that development of integrated care for CYP at scale in England has the potential to dramatically reduce or even reverse these forecast increases, reducing strain in the system whilst improving outcomes for CYP and family and young people’s experience of care. Development of integrated care systems for CYP was identified as a priority by the NHS England Long Term Plan in early 2019.^13^ However integrated care for CYP in England starts from a low baseline in many areas, with great variation in the degree to which ICSs include provision for CYP.^30^ The many harms brought by the COVID-19 pandemic have included delays to Long Term Plan programmes including integrated care for CYP. Yet the pandemic has also driven take-up in innovation (eg. digital innovations) and suggested that non-essential activity can be reduced without necessarily worsening health outcomes,^16^ supporting policy efforts to build upon beneficial changes made during the pandemic.

## Data Availability

Data not available

## Acknowledgements

Members of the Paediatrics 2040 team, both Commissions and members of the RCPCH staff.

DH is supported by the National Institute for Health Research (NIHR) through the National School for Public Health Research Programme and the Applied Health Research (ARC) programme for North West London. The views expressed in this publication are those of the authors and not necessarily those of the of any funding bodies or institutions mentioned above.

## Data sharing

All observed data used here are available from Hospital Episode Statistics (HES) through NHS Digital.

