## Appendix for "Recent and forecast post-COVID trends in hospital activity in England amongst 0 to 24 year olds: analyses using routine hospital administrative data"

#### Web extra material

**Appendix Table 1. Cause categories used in this study mapped to Global Burden of Disease (GBD) level 3 causes**

| CAUSE | GBD Level 3 CAUSES |
|---|---|
| Acne | Acne vulgaris |
| Adverse effects | Allergy / adverse effect |
| Anxiety & Depression | Anxiety disorders |
| | Bipolar disorder |
| | Depressive disorders |
| Arthritis | Gout |
| | Osteoarthritis |
| | Other musculoskeletal disorders |
| | Rheumatoid arthritis |
| Cardiovascular | Aortic aneurysm |
| | Atrial fibrillation and flutter |
| | Cardiomyopathy and myocarditis |
| | Endocarditis |
| | Hypertension |
| | Hypertensive heart disease |
| | Ischemic heart disease |
| | Non-rheumatic valvular heart disease |
| | Other cardiovascular and circulatory diseases |
| | Peripheral artery disease |
| | Rheumatic heart disease |
| | Stroke |
| Chronic renal | Acute glomerulonephritis |
| | Chronic kidney disease |
| | Pulmonary embolism |
| | Urinary diseases and male infertility |
| Chronic respiratory | Aspiration pneumonia |
| | Chronic obstructive pulmonary disease |
| | Interstitial lung disease and pulmonary sarcoidosis |
| | Other chronic respiratory diseases |
| | Pneumoconiosis |
| CNS cancer | Brain and nervous system cancer |
| Conduct | Conduct disorder |
| Congenital | Congenital birth defects |
| Diabetes | Diabetes mellitus |
| | DKA |
| Diarrhoea | Diarrheal diseases |
| | Other intestinal infectious diseases |
| Eating disorders | Eating disorders |
| Endocrine | Endocrine, metabolic, blood, and immune disorders |
| Epilepsy | Convulsions |
| | Epilepsy |
| Fever and nonspecific viral infections | Other unspecified infection only viral unspecified |
| Foreign body | Foreign body |

| | |
|---|---|
| Gastrointestinal | Appendicitis |
| | Cirrhosis and other chronic liver diseases |
| | Gallbladder and biliary diseases |
| | Inflammatory bowel disease |
| | Inguinal, femoral, and abdominal hernia |
| | Other digestive diseases |
| | Pancreatitis |
| | Paralytic ileus and intestinal obstruction |
| | Upper digestive system diseases |
| | Vascular intestinal disorders |
| Gynaecological | Gynecological diseases |
| Haematological cancer | Hodgkin lymphoma |
| | Leukemia |
| | Multiple myeloma |
| | Non-Hodgkin lymphoma |
| Haematology non-malignant | Hemoglobinopathies and hemolytic anemias |
| Headache | Headache disorders |
| Hearing | Disorders of the ear |
| HIV/AIDS | HIV/AIDS |
| Injuries | Injury |
| | Poisonings |
| Learning difficulties | Idiopathic developmental intellectual disability |
| Lower Respiratory Infection | Lower respiratory infections |
| Maternal | Maternal disorders |
| Meningitis | Encephalitis |
| | Meningitis |
| Neonatal / prematurity | Neonatal disorders |
| Neurodevelopmental | Attention-deficit/hyperactivity disorder |
| | Autism spectrum disorders |
| | Other mental disorders |
| Neurological | Alzheimer's disease and other dementias |
| | Motor neuron disease |
| | Multiple sclerosis |
| | Other neurological disorders |
| | Parkinson's disease |
| | Schizophrenia |
| Non-specific symptoms | Abnormal finding on examination |
| | Abnormal investigation |
| | Abnormal mobility |
| | Abnormal movements |
| | Follow up / general examination |
| | Observation |
| | Other |
| | Sign: Lymphadenopathy |
| | Sign: Oedema |
| | Symptom : abnormal sensations |
| | Symptom : Nausea and vomiting |
| | Symptom: abdominal and pelvic pain |

| | |
|---|---|
| | Symptom: bleeding |
| | Symptom: Other |
| Nutrition | Dietary iron deficiency |
| | Other nutritional deficiencies |
| | Protein-energy malnutrition |
| | Vitamin A deficiency |
| Oral | Oral disorders |
| Other bacterial infections | Bacterial skin diseases |
| | Other unspecified infection excluding viral unspecified |
| | Otitis media |
| | Varicella and herpes zoster |
| Other cancer | Bladder cancer |
| | Breast cancer |
| | Cervical cancer |
| | Colon and rectum cancer |
| | Esophageal cancer |
| | Gallbladder and biliary tract cancer |
| | Larynx cancer |
| | Lip and oral cavity cancer |
| | Liver cancer |
| | Malignant skin melanoma |
| | Mesothelioma |
| | Nasopharynx cancer |
| | Non-melanoma skin cancer |
| | Other malignant neoplasms |
| | Other neoplasms |
| | Other pharynx cancer |
| | Ovarian cancer |
| | Pancreatic cancer |
| | Prostate cancer |
| | Stomach cancer |
| | Testicular cancer |
| | Thyroid cancer |
| | Tracheal, bronchus, and lung cancer |
| | Uterine cancer |
| Pain | Pain, other |
| Prevention & immunisation | Procedure : Immunization |
| | Screening |
| | Treatment : prophylaxis |
| Rare infections | Acute hepatitis |
| | African trypanosomiasis |
| | Chagas disease |
| | Cystic echinococcosis |
| | Cysticercosis |
| | Dengue |
| | Diphtheria |
| | Ebola |
| | Food-borne trematodiases |

| | |
|---|---|
| | Guinea worm disease |
| | Intestinal nematode infections |
| | Invasive Non-typhoidal Salmonella (iNTS) |
| | Leishmaniasis |
| | Leprosy |
| | Lymphatic filariasis |
| | Malaria |
| | Measles |
| | Other neglected tropical diseases |
| | Rabies |
| | Schistosomiasis |
| | Tetanus |
| | Trachoma |
| | Tuberculosis |
| | Typhoid and paratyphoid |
| | Whooping cough |
| | Yellow fever |
| | Zika virus |
| Renal including Wilms | Kidney cancer |
| SIDS | Sudden infant death syndrome |
| Skin | Alopecia areata |
| | Decubitus ulcer |
| | Dermatitis |
| | Other skin and subcutaneous diseases |
| | Pruritus |
| | Psoriasis |
| | Scabies |
| | Urticaria |
| STD | Sexually transmitted infections excluding HIV |
| Substance use | Alcohol use disorders |
| | Drug use disorders |
| URTI | Upper respiratory infections |
| Vision | Opthalmological disorders |
| Wheeze/Asthma | Asthma |
| | Symptom: cough, wheeze, shortness of breath, other breathing problem |

### Appendix Table 2. Attendance and admission rates in 2007 and 2017 and change in each rate from 2007 to 2017, by region and age-group

**A&E Attendance**

| | | infants | 1 to 4 | 5 to 9 | 10 to 14 | 15 to 19 | 20 to 24 |
|---|---|---|---|---|---|---|---|
| England | 2007 | 47,934 | 33,372 | 18,277 | 22,895 | 28,193 | 29,700 |
| | 2017 | 87,412 | 54,686 | 29,806 | 33,553 | 38,071 | 43,608 |
| | | 82.4% | 63.9% | 63.1% | 46.6% | 35.0% | 46.8% |
| East Midlands | 2007 | 39,301 | 29,559 | 16,396 | 22,240 | 27,237 | 27,229 |
| | 2017 | 79,191 | 52,005 | 27,909 | 31,161 | 37,092 | 40,009 |
| | | 101.5% | 75.9% | 70.2% | 40.1% | 36.2% | 46.9% |
| East of England | 2007 | 33,458 | 24,888 | 14,374 | 19,902 | 23,844 | 25,535 |
| | 2017 | 68,726 | 43,156 | 24,083 | 29,678 | 32,360 | 37,119 |
| | | 105.4% | 73.4% | 67.5% | 49.1% | 35.7% | 45.4% |
| London | 2007 | 74,049 | 46,750 | 24,650 | 25,378 | 31,534 | 33,238 |
| | 2017 | 112,461 | 61,745 | 33,491 | 33,439 | 40,422 | 54,083 |
| | | 51.9% | 32.1% | 35.9% | 31.8% | 28.2% | 62.7% |
| North East | 2007 | 54,093 | 40,615 | 21,612 | 26,466 | 32,756 | 32,492 |
| | 2017 | 132,280 | 79,164 | 40,841 | 43,401 | 49,098 | 51,502 |
| | | 144.5% | 94.9% | 89.0% | 64.0% | 49.9% | 58.5% |
| North West | 2007 | 58,516 | 40,813 | 22,116 | 28,008 | 35,458 | 38,695 |
| | 2017 | 107,614 | 68,772 | 35,892 | 39,455 | 44,243 | 50,009 |
| | | 83.9% | 68.5% | 62.3% | 40.9% | 24.8% | 29.2% |
| South East | 2007 | 31,642 | 23,982 | 13,988 | 19,128 | 22,696 | 23,583 |
| | 2017 | 71,316 | 49,041 | 27,262 | 31,860 | 35,257 | 40,041 |
| | | 125.4% | 104.5% | 94.9% | 66.6% | 55.3% | 69.8% |
| South West | 2007 | 27,574 | 21,677 | 13,093 | 18,049 | 21,496 | 21,544 |
| | 2017 | 59,416 | 43,779 | 26,740 | 34,006 | 35,966 | 38,235 |
| | | 115.5% | 102.0% | 104.2% | 88.4% | 67.3% | 77.5% |
| West Midlands | 2007 | 57,916 | 40,862 | 22,299 | 27,692 | 34,827 | 38,113 |
| | 2017 | 83,305 | 52,284 | 28,716 | 32,410 | 35,638 | 39,326 |
| | | 43.8% | 28.0% | 28.8% | 17.0% | 2.3% | 3.2% |
| Yorkshire and Humber | 2007 | 37,138 | 26,694 | 15,239 | 20,200 | 24,673 | 23,965 |
| | 2017 | 73,532 | 49,088 | 26,905 | 31,537 | 37,672 | 39,729 |
| | | 98.0% | 83.9% | 76.6% | 56.1% | 52.7% | 65.8% |

**All Admissions**

| | | infants | 1 to 4 | 5 to 9 | 10 to 14 | 15 to 19 | 20 to 24 |
|---|---|---|---|---|---|---|---|
| England | 2007 | 33,279 | 15,239 | 8,637 | 7,753 | 13,440 | 20,135 |
| | 2017 | 38,574 | 16,103 | 8,918 | 8,466 | 12,207 | 17,980 |
| | | 15.9% | 5.7% | 3.3% | 9.2% | -9.2% | -10.7% |
| East Midlands | 2007 | 32,209 | 14,391 | 7,657 | 6,948 | 13,676 | 20,782 |
| | 2017 | 35,604 | 14,384 | 7,818 | 7,295 | 11,783 | 18,611 |
| | | 10.5% | -0.1% | 2.1% | 5.0% | -13.8% | -10.4% |
| East of England | 2007 | 25,644 | 11,767 | 6,805 | 6,511 | 11,597 | 18,880 |
| | 2017 | 37,491 | 14,065 | 7,435 | 7,833 | 11,904 | 18,538 |
| | | 46.2% | 19.5% | 9.3% | 20.3% | 2.6% | -1.8% |
| London | 2007 | 23,468 | 11,999 | 8,092 | 7,099 | 12,041 | 17,683 |
| | 2017 | 29,510 | 13,444 | 8,905 | 8,558 | 11,360 | 16,563 |
| | | 25.7% | 12.0% | 10.1% | 20.6% | -5.7% | -6.3% |
| North East | 2007 | 47,394 | 21,611 | 11,515 | 9,605 | 16,584 | 21,624 |
| | 2017 | 49,847 | 21,150 | 10,871 | 9,780 | 13,123 | 17,030 |
| | | 5.2% | -2.1% | -5.6% | 1.8% | -20.9% | -21.2% |
| North West | 2007 | 45,285 | 19,915 | 10,621 | 9,331 | 16,185 | 23,807 |
| | 2017 | 51,791 | 21,439 | 11,301 | 10,615 | 14,088 | 20,289 |
| | | 14.4% | 7.7% | 6.4% | 13.8% | -13.0% | -14.8% |
| South East | 2007 | 32,811 | 14,774 | 7,946 | 7,265 | 11,116 | 16,592 |
| | 2017 | 37,151 | 15,340 | 7,960 | 7,556 | 11,044 | 15,678 |
| | | 13.2% | 3.8% | 0.2% | 4.0% | -0.6% | -5.5% |
| South West | 2007 | 31,087 | 15,450 | 9,174 | 8,680 | 13,791 | 20,413 |
| | 2017 | 39,225 | 15,614 | 7,918 | 7,535 | 12,139 | 18,493 |
| | | 26.2% | 1.1% | -13.7% | -13.2% | -12.0% | -9.4% |
| West Midlands | 2007 | 38,411 | 16,155 | 8,281 | 7,608 | 13,519 | 22,141 |
| | 2017 | 41,367 | 17,114 | 9,055 | 8,836 | 12,550 | 18,141 |
| | | 7.7% | 5.9% | 9.4% | 16.2% | -7.2% | -18.1% |
| Yorkshire and Humber | 2007 | 35,710 | 15,712 | 9,365 | 7,736 | 14,884 | 21,858 |
| | 2017 | 36,079 | 16,082 | 9,960 | 8,607 | 12,858 | 19,276 |
| | | 1.0% | 2.4% | 6.4% | 11.3% | -13.6% | -11.8% |

**Emergency Admissions**

| | | infants | 1 to 4 | 5 to 9 | 10 to 14 | 15 to 19 | 20 to 24 |
|---|---|---|---|---|---|---|---|
| England | 2007 | 22,482 | 8,221 | 3,119 | 3,171 | 4,880 | 5,603 |
| | 2017 | 29,658 | 10,120 | 3,839 | 3,726 | 5,321 | 6,276 |
| | | 31.9% | 23.1% | 23.1% | 17.5% | 9.0% | 12.0% |
| East Midlands | 2007 | 23,109 | 8,041 | 2,956 | 2,998 | 4,746 | 5,490 |
| | 2017 | 27,267 | 9,158 | 3,459 | 3,282 | 5,066 | 5,941 |
| | | 18.0% | 13.9% | 17.0% | 9.5% | 6.7% | 8.2% |
| East of England | 2007 | 16,437 | 5,967 | 2,449 | 2,634 | 3,914 | 4,614 |
| | 2017 | 28,399 | 8,228 | 3,295 | 3,243 | 5,030 | 6,282 |
| | | 72.8% | 37.9% | 34.6% | 23.1% | 28.5% | 36.2% |
| London | 2007 | 14,196 | 5,628 | 2,583 | 2,666 | 4,383 | 4,949 |
| | 2017 | 19,743 | 7,311 | 3,137 | 3,110 | 4,370 | 5,376 |
| | | 39.1% | 29.9% | 21.4% | 16.6% | -0.3% | 8.6% |
| North East | 2007 | 32,737 | 11,886 | 4,226 | 4,043 | 5,970 | 6,573 |
| | 2017 | 39,449 | 13,311 | 4,687 | 4,509 | 5,961 | 6,305 |
| | | 20.5% | 12.0% | 10.9% | 11.5% | -0.2% | -4.1% |
| North West | 2007 | 32,938 | 11,785 | 4,093 | 4,090 | 6,168 | 7,056 |
| | 2017 | 42,192 | 14,732 | 5,148 | 4,889 | 6,075 | 7,207 |
| | | 28.1% | 25.0% | 25.8% | 19.5% | -1.5% | 2.1% |
| South East | 2007 | 21,060 | 7,477 | 2,787 | 2,846 | 4,448 | 5,111 |
| | 2017 | 29,192 | 9,794 | 3,685 | 3,491 | 5,249 | 6,213 |
| | | 38.6% | 31.0% | 32.2% | 22.7% | 18.0% | 21.6% |
| South West | 2007 | 20,811 | 7,987 | 3,001 | 3,180 | 4,837 | 5,556 |
| | 2017 | 32,244 | 10,667 | 3,780 | 3,858 | 5,775 | 6,501 |
| | | 54.9% | 33.6% | 25.9% | 21.3% | 19.4% | 17.0% |
| West Midlands | 2007 | 25,612 | 9,297 | 3,330 | 3,133 | 4,689 | 5,707 |
| | 2017 | 32,033 | 11,503 | 4,385 | 4,151 | 5,556 | 6,574 |
| | | 25.1% | 23.7% | 31.7% | 32.5% | 18.5% | 15.2% |
| Yorkshire and Humber | 2007 | 25,979 | 9,099 | 3,436 | 3,509 | 5,264 | 5,886 |
| | 2017 | 27,373 | 9,291 | 3,610 | 3,546 | 5,389 | 6,291 |
| | | 5.4% | 2.1% | 5.0% | 1.1% | 2.4% | 6.9% |

**Elective admissions**

| | | infants | 1 to 4 | 5 to 9 | 10 to 14 | 15 to 19 | 20 to 24 |
|---|---|---|---|---|---|---|---|
| England | 2007 | 2,174 | 1,635 | 1,334 | 1,277 | 1,509 | 1,528 |
| | 2017 | 1,973 | 1,381 | 905 | 961 | 1,034 | 971 |
| | | -9.3% | -15.5% | -32.2% | -24.7% | -31.5% | -36.4% |
| East Midlands | 2007 | 1,897 | 1,658 | 1,340 | 1,122 | 1,486 | 1,537 |
| | 2017 | 1,825 | 1,297 | 825 | 763 | 932 | 984 |
| | | -3.8% | -21.7% | -38.5% | -32.0% | -37.2% | -36.0% |
| East of England | 2007 | 2,483 | 1,658 | 1,227 | 1,239 | 1,547 | 1,569 |
| | 2017 | 2,133 | 1,630 | 953 | 980 | 1,095 | 1,106 |
| | | -14.1% | -1.7% | -22.4% | -20.9% | -29.2% | -29.5% |
| London | 2007 | 2,121 | 1,474 | 1,076 | 1,049 | 1,208 | 1,162 |
| | 2017 | 1,987 | 1,296 | 823 | 891 | 931 | 911 |
| | | -6.3% | -12.0% | -23.5% | -15.0% | -22.9% | -21.6% |
| North East | 2007 | 1,944 | 1,860 | 1,751 | 1,628 | 2,076 | 2,023 |
| | 2017 | 2,283 | 2,037 | 1,569 | 1,490 | 1,208 | 1,012 |
| | | 17.4% | 9.5% | -10.4% | -8.5% | -41.8% | -50.0% |
| North West | 2007 | 2,734 | 1,913 | 1,403 | 1,343 | 1,610 | 1,743 |
| | 2017 | 2,180 | 1,460 | 962 | 1,102 | 965 | 937 |
| | | -20.3% | -23.7% | -31.4% | -17.9% | -40.0% | -46.3% |
| South East | 2007 | 1,858 | 1,533 | 1,292 | 1,215 | 1,360 | 1,452 |
| | 2017 | 2,055 | 1,336 | 804 | 840 | 1,003 | 952 |
| | | 10.6% | -12.8% | -37.7% | -30.8% | -26.2% | -34.4% |
| South West | 2007 | 2,554 | 1,901 | 1,708 | 1,698 | 1,723 | 1,656 |
| | 2017 | 1,743 | 1,204 | 740 | 788 | 1,008 | 997 |
| | | -31.7% | -36.7% | -56.6% | -53.6% | -41.5% | -39.8% |
| West Midlands | 2007 | 1,850 | 1,311 | 1,245 | 1,231 | 1,436 | 1,414 |
| | 2017 | 1,746 | 1,236 | 952 | 1,172 | 1,319 | 967 |
| | | -5.6% | -5.7% | -23.5% | -4.8% | -8.1% | -31.6% |
| Yorkshire and Humber | 2007 | 2,036 | 1,676 | 1,370 | 1,256 | 1,575 | 1,654 |
| | 2017 | 1,798 | 1,329 | 954 | 932 | 975 | 957 |
| | | -11.7% | -20.7% | -30.4% | -25.8% | -38.1% | -42.2% |

**Appendix Table 3. Heat-map of proportional change from 2007 to 2017 in main causes of emergency admissions by age**

| | 7 days to 1 year | | 1 to 4 | | 5 to 9 | | 10 to 14 | | 15 to 19 | | 20 to 24 | |
|---|---|---|---|---|---|---|---|---|---|---|---|---|
| Cause | 2012 | 2017 | 2012 | 2017 | 2012 | 2017 | 2012 | 2017 | 2012 | 2017 | 2012 | 2017 |
| Arthritis | | | 0.2% | 13.5% | 4.2% | 10.4% | 11.1% | 35.0% | -7.4% | 13.5% | -3.6% | 17.8% |
| Cardiovascular | | | | | 81.5% | 167.0% | 75.9% | 136.0% | | | | |
| Chronic renal | 16.6% | 24.2% | 2.1% | 14.4% | 11.7% | 32.1% | 22.4% | 52.6% | 16.5% | 35.9% | 32.2% | 45.8% |
| Common infections | 42.8% | 56.9% | 142.5% | 208.2% | 87.5% | 135.7% | 23.2% | 60.5% | 37.0% | 141.6% | -1.9% | 61.9% |
| Congenital | 3.6% | 27.8% | | | | | | | | | | |
| Diabetes | | | | | -49.7% | -71.9% | -2.0% | -10.6% | 10.0% | 12.1% | 52.4% | 103.2% |
| Diarrhoea | 42.6% | -8.8% | 41.2% | -5.7% | 79.7% | 122.3% | 146.7% | 189.4% | | | | |
| Epilepsy | -15.9% | -24.5% | -21.1% | -34.0% | -2.1% | 10.3% | -5.2% | 6.2% | 4.4% | 9.0% | -74.0% | -100.0% |
| Fever & unspecified infections | 51.9% | 66.9% | 162.1% | 225.6% | 105.1% | 142.1% | 38.2% | 82.0% | 13.7% | 92.3% | 72.2% | 209.1% |
| Foreign body | | | -61.2% | -100.0% | -100.0% | -100.0% | | | | | | |
| GI disorders | -36.3% | -18.6% | -72.4% | -70.0% | -25.7% | -15.5% | -15.7% | -15.0% | -21.1% | -25.5% | 3.7% | -2.0% |
| Gynaecological | | | | | -100.0% | -100.0% | 20.7% | 35.3% | -1.9% | 6.1% | 3.1% | 4.9% |
| Haem non-malig | | | | | 91.9% | -10.0% | 53.1% | 167.4% | 3.0% | 5.2% | 190.4% | 39.9% |
| Headache | | | | | | | 98.6% | 447.4% | | | | |
| Injuries | 3.1% | 24.7% | -0.1% | -5.3% | -14.1% | -15.3% | -19.4% | -12.9% | -22.6% | -13.8% | -13.0% | -18.4% |
| Lower Resp Infection | 26.2% | 71.2% | 39.8% | 76.0% | 55.7% | 69.7% | 35.0% | 96.3% | 56.0% | 148.2% | 28.2% | 92.8% |
| Maternal | | | | | | | | | -25.8% | -33.2% | -7.1% | -8.3% |
| Neonatal / prematurity | 35.5% | 90.3% | | | | | | | | | | |
| Neurodevelopmental | 12.2% | 55.7% | -1.5% | 22.8% | 23.9% | 49.0% | 38.1% | 116.8% | 20.2% | 84.1% | 16.4% | 43.9% |
| Non-specific sympt | -12.4% | 46.3% | -45.2% | -71.8% | -10.1% | -0.4% | -4.3% | -3.1% | 0.4% | -7.3% | 7.8% | 12.0% |
| Oral | | | | | | | | | | | | |
| Pain | | | | | | | 11.5% | 38.1% | -5.2% | 14.7% | 2.0% | 17.9% |
| Skin | -17.0% | -4.0% | -9.2% | -8.0% | 4.5% | 11.0% | 4.3% | 33.4% | | | | |
| Subs use | | | | | | | -100.0% | -100.0% | -47.6% | -89.7% | -100.0% | -12.5% |
| URTI | -4.1% | 30.8% | -1.6% | 32.4% | 17.1% | 38.0% | 4.6% | 43.7% | 4.1% | 66.6% | 18.4% | 83.6% |
| Wheeze/Asthma | -32.1% | -37.3% | -33.8% | -53.2% | 13.9% | 5.7% | 16.5% | 34.6% | 9.6% | 28.3% | 3.9% | 20.9% |

The heat-map shows proportional change (increase or decrease) from 2007 to 2017 in the main causes of emergency admissions by age. Colours are graded for change, with reductions are shown in blue and increases in orange.

**Appendix Table 4. Outpatient attendance rates and change in thse rates from 2007 to 2017, by age**

Outpatient attendances and change from 2007 to 2017, by age

| | infants | 1 to 4 | 5 to 9 | 10 to 14 | 15 to 19 | 20 to 24 |
|---|---|---|---|---|---|---|
| 2007 | 80,550 | 54,704 | 49,710 | 53,382 | 52,501 | 47,234 |
| | 0% | 0% | 0% | 0% | 0% | 0% |
| 2012 | 104,784 | 66,362 | 62,180 | 64,619 | 64,787 | 57,702 |
| | 30% | 21% | 25% | 21% | 23% | 22% |
| 2017 | 138,690 | 73,322 | 68,535 | 74,774 | 77,177 | 67,761 |
| | 72% | 34% | 38% | 40% | 47% | 43% |

**Appendix Table 5. Projection model parameters for emergency activity from 2018 to 2040**

| | | Emergency Attendances | | | | | Emergency Admissions | | | | |
|---|---|---|---|---|---|---|---|---|---|---|---|
| | | Mean | Std. Dev. | Median | [95% Credible interval] | | Mean | Std. Dev. | Median | [95% Credible interval] | |
| Infant | Year | 10636.9 | 9.223834 | 10637.78 | 10620.12 | 10650.91 | 3813.726 | 900.5417 | 3727.153 | 3175.902 | 4462.29 |
| | Population | 2.050419 | 0.0001478 | 2.050414 | 2.050151 | 2.050747 | 0.2565725 | 0.0064399 | 0.2571793 | 0.2519188 | 0.2612019 |
| | % minority ethnicity | 7511433 | 14.25107 | 7511434 | 7511407 | 7511455 | 1254867 | 771.6668 | 1254785 | 1254315 | 1255424 |
| | Child poverty | -485217.3 | 2.717164 | -485216.9 | -485222.3 | -485213.2 | 87969.3 | 210.3195 | 87962.4 | 87820.56 | 88118.96 |
| | Intercept | -2636243 | 22.86926 | -2636245 | -2636277 | -2636201 | -345438.9 | 452.4112 | -345430.8 | -345760 | -345116.8 |
| 1-4y | Year | 2205.254 | 1501.283 | 1299.867 | 1154.463 | 4303.924 | 1682.796 | 20.22894 | 1688.918 | 1659.662 | 1699.258 |
| | Population | 1.666834 | 0.0023285 | 1.667568 | 1.663819 | 1.668605 | 0.1842126 | 0.0001366 | 0.1842254 | 0.1840648 | 0.1843608 |
| | % minority ethnicity | 2.30E+07 | 3309.751 | 2.30E+07 | 2.30E+07 | 2.30E+07 | 1449871 | 629.4623 | 1449751 | 1449309 | 1450552 |
| | Child poverty | 73953.49 | 2442.531 | 75344.06 | 70669 | 75603.48 | 433606.8 | 451.3354 | 433503.5 | 433216.1 | 434100.9 |
| | Intercept | -8799780 | 728.3335 | -8799442 | -8801105 | -8798831 | -730885.5 | 462.4402 | -730685.9 | -731416.4 | -730556.3 |
| 5-9y | Year | 53430.58 | 8.34E+09 | 53430.58 | 53430.58 | 53430.58 | 3858.071 | 62.16423 | 3876.711 | 3784.476 | 3900.845 |
| | Population | -0.0308466 | 5.04E+11 | -0.0308466 | -0.0308466 | -0.0308466 | 0.0254181 | 0.0359466 | 0.0253517 | 3.47E-17 | 0.0509065 |
| | % minority ethnicity | | | | | | 506638.5 | 715387.2 | 506729.9 | 607.1809 | 1012494 |
| | Child poverty | -690899.4 | 8.35E+09 | -690899.4 | -690899.4 | -690899.4 | 130483.7 | 305.6099 | 130499.1 | 130242.4 | 130699.6 |
| | Intercept | 837361.1 | 9.73E+09 | 837361.1 | 837361.1 | 837361.1 | -107166.8 | 39.62325 | -107153.4 | -107255 | -107100.3 |
| 10-14y | Year | 14.74726 | 15.92215 | 8.421554 | 1.798167 | 28.49108 | 2186.915 | 32.8217 | 2186.908 | 2163.703 | 2210.14 |
| | Population | 7.65E-06 | 0.0000227 | 3.87E-06 | 2.49E-07 | 0.0000164 | 0.0306183 | 0.0106741 | 0.0306085 | 0.0230695 | 0.0381734 |
| | % minority ethnicity | | | | | | | | | | |
| | Child poverty | -1137469 | 95.54712 | -1137472 | -1137518 | -1137409 | 79325.94 | 112201.7 | 79326.1 | -12.83132 | 158664.6 |
| | Intercept | 285166.5 | 74.77992 | 285161.6 | 285114.5 | 285206.7 | -23982.11 | 67.41228 | -23982.12 | -24029.78 | -23934.42 |
| 15-19y | Year | 54620.6 | 624.3802 | 54356.33 | 54163.02 | 55473.15 | 1070.396 | 85.27146 | 1070.825 | 970.2816 | 1278.443 |
| | Population | 1.550387 | 0.0193988 | 1.548414 | 1.531574 | 1.570929 | 0.0361284 | 0.0073063 | 0.0350253 | 0.0310683 | 0.0459959 |
| | % minority ethnicity | | | | | | | | | | |
| | Child poverty | -148137 | 257570.2 | -38.2672 | -445554.4 | 2214.518 | 102892 | 141272.9 | 103260 | 1042.139 | 202786.2 |

| | | | | | | | | | | |
|---|---|---|---|---|---|---|---|---|---|---|
| Intercept | -4164946 | 36385.78 | -4144860 | -4210126 | -4143056 | 7931.253 | 18511.95 | -3686.565 | -4675.807 | 26918.92 |

#### Appendix Table 6. ED attendances, Emergency Admissions and Total Admissions from 2017 to 2040 in scenarios of stable, increasing and decreasing poverty and COVID-19 scenarios.

| | | ED attendances | | | | | | Emergency Admissions | | | | | | All Admissions | | | | | |
|---|---|---|---|---|---|---|---|---|---|---|---|---|---|---|---|---|---|---|---|
| | | 2017 | 2020 | 2025 | 2030 | 2035 | 2040 | 2017 | 2020 | 2025 | 2030 | 2035 | 2040 | 2017 | 2020 | 2025 | 2030 | 2035 | 2040 |
| infant | Stable poverty projection | 87,072 | 109,821 | 139,421 | 167,606 | 192,079 | 212,787 | 30,169 | 33,068 | 38,294 | 43,297 | 47,367 | 50,801 | 35,684 | 38,583 | 43,809 | 48,812 | 52,882 | 56,315 |
| | 99% CI lower | 86,423 | 107,935 | 136,416 | 163,517 | 186,845 | 206,466 | 29,666 | 32,354 | 37,149 | 41,728 | 45,422 | 48,499 | 35,102 | 37,790 | 42,585 | 47,164 | 50,858 | 53,935 |
| | 99% CI upper | 87,498 | 111,385 | 141,911 | 170,990 | 196,438 | 218,071 | 30,731 | 33,871 | 39,589 | 45,072 | 49,570 | 53,409 | 36,326 | 39,466 | 45,183 | 50,667 | 55,165 | 59,003 |
| | % change - poverty stable | 0% | 26% | 60% | 92% | 121% | 144% | 0% | 10% | 27% | 44% | 57% | 68% | 0% | 8% | 23% | 37% | 48% | 58% |
| | Decreasing poverty projection | 91,336 | 115,304 | 149,914 | 182,825 | 210,276 | 232,998 | 30,706 | 33,354 | 38,839 | 44,360 | 48,928 | 52,837 | 36,221 | 38,868 | 44,354 | 49,875 | 54,443 | 58,352 |
| | 99%CI lower | 90,818 | 114,307 | 147,370 | 178,761 | 205,322 | 227,621 | 30,148 | 32,674 | 37,738 | 42,913 | 47,179 | 50,812 | 35,584 | 38,110 | 43,174 | 48,349 | 52,615 | 56,248 |
| | 99%CI upper | 91,906 | 116,275 | 151,913 | 185,821 | 213,879 | 236,980 | 31,354 | 33,999 | 39,877 | 45,935 | 50,976 | 55,323 | 36,949 | 39,594 | 45,472 | 51,529 | 56,570 | 60,918 |
| | % change - decreasing poverty | 0% | 26% | 64% | 100% | 130% | 155% | 0% | 9% | 26% | 44% | 59% | 72% | 0% | 7% | 22% | 38% | 50% | 61% |
| | Increasing poverty projection | 89,148 | 110,900 | 142,054 | 172,122 | 198,339 | 220,696 | 30,716 | 34,631 | 40,505 | 46,006 | 50,506 | 54,345 | 36,231 | 40,146 | 46,020 | 51,521 | 56,021 | 59,860 |
| | 99%CI lower | 87,492 | 107,184 | 137,006 | 166,144 | 191,719 | 213,473 | 30,687 | 34,606 | 40,495 | 45,993 | 50,481 | 54,309 | 36,123 | 40,042 | 45,931 | 51,429 | 55,917 | 59,745 |
| | 99%CI upper | 90,862 | 114,682 | 147,189 | 178,214 | 205,095 | 228,076 | 30,751 | 34,663 | 40,523 | 46,013 | 50,524 | 54,375 | 36,346 | 40,258 | 46,118 | 51,608 | 56,119 | 59,969 |
| | % change - increasing poverty | 0% | 24% | 59% | 93% | 122% | 148% | 0% | 13% | 32% | 50% | 64% | 77% | 0% | 11% | 27% | 42% | 55% | 65% |
| | COVID shock | | 54,910 | 139,421 | 167,606 | 192,079 | 212,787 | | 24,801 | 38,294 | 43,297 | 47,367 | 50,801 | | 27,558 | 43,809 | 48,812 | 52,882 | 56,315 |
| | Integrated care - moderate | | 54,910 | 106,657 | 128,219 | 146,941 | 162,782 | | 24,801 | 29,295 | 33,122 | 36,236 | 38,862 | | 27,558 | 34,810 | 38,637 | 41,750 | 44,377 |
| | Integrated care - high | | 54,910 | 73,893 | 88,831 | 101,802 | 112,777 | | 24,801 | 20,296 | 22,947 | 25,104 | 26,924 | | 27,558 | 25,811 | 28,462 | 30,619 | 32,439 |
| 1 to 4 | Stable poverty projection | 55,457 | 65,495 | 85,179 | 103,914 | 121,017 | 135,035 | 10,104 | 10,856 | 12,331 | 13,719 | 15,015 | 16,117 | 15,327 | 16,079 | 17,554 | 18,942 | 20,237 | 21,339 |
| | 99% CI lower | 55,032 | 64,818 | 84,189 | 102,517 | 119,319 | 133,080 | 10,056 | 10,774 | 12,197 | 13,531 | 14,778 | 15,834 | 15,242 | 15,960 | 17,383 | 18,717 | 19,963 | 21,020 |
| | 99% CI upper | 55,898 | 66,233 | 86,288 | 105,503 | 122,960 | 137,260 | 10,120 | 10,882 | 12,377 | 13,785 | 15,099 | 16,218 | 15,380 | 16,141 | 17,636 | 19,044 | 20,359 | 21,478 |
| | % change - poverty stable | 0% | 18% | 54% | 87% | 118% | 143% | 0% | 7% | 22% | 36% | 49% | 60% | 0% | 5% | 15% | 24% | 32% | 39% |
| | Decreasing poverty projection | 54,874 | 63,681 | 82,004 | 99,491 | 115,585 | 128,847 | 10,268 | 10,085 | 11,271 | 12,835 | 14,308 | 15,579 | 15,491 | 15,307 | 16,494 | 18,058 | 19,531 | 20,802 |
| | 99%CI lower | 54,433 | 62,653 | 80,153 | 96,887 | 112,327 | 125,031 | 10,199 | 9,978 | 11,110 | 12,612 | 14,033 | 15,248 | 15,385 | 15,163 | 16,295 | 17,798 | 19,218 | 20,434 |
| | 99%CI upper | 55,357 | 64,128 | 82,850 | 100,609 | 116,927 | 130,342 | 10,417 | 10,324 | 11,652 | 13,353 | 14,955 | 16,343 | 15,677 | 15,584 | 16,912 | 18,613 | 20,215 | 21,603 |
| | % change - decreasing poverty | 0% | 16% | 49% | 81% | 111% | 135% | 0% | -2% | 10% | 25% | 39% | 52% | 0% | -1% | 6% | 17% | 26% | 34% |
| | Increasing poverty projection | 54,857 | 64,447 | 83,274 | 101,031 | 117,341 | 130,745 | 10,134 | 11,855 | 13,499 | 14,898 | 16,189 | 17,280 | 15,356 | 17,078 | 18,722 | 20,120 | 21,412 | 22,502 |
| | 99%CI lower | 54,842 | 64,434 | 83,263 | 101,024 | 117,336 | 130,741 | 10,133 | 11,855 | 13,499 | 14,897 | 16,189 | 17,279 | 15,319 | 17,040 | 18,684 | 20,083 | 21,374 | 22,465 |
| | 99%CI upper | 54,873 | 64,460 | 83,285 | 101,039 | 117,347 | 130,750 | 10,134 | 11,856 | 13,499 | 14,898 | 16,190 | 17,280 | 15,394 | 17,115 | 18,759 | 20,158 | 21,449 | 22,540 |
| | % change - increasing poverty | 0% | 17% | 52% | 84% | 114% | 138% | 0% | 17% | 33% | 47% | 60% | 71% | 0% | 11% | 22% | 31% | 39% | 47% |
| | COVID shock | | 32,748 | 85,179 | 103,914 | 121,017 | 135,035 | | 8,142 | 12,331 | 13,719 | 15,015 | 16,117 | | 10,754 | 17,554 | 18,942 | 20,237 | 21,339 |
| | Integrated care - moderate | | 32,748 | 57,070 | 69,623 | 81,081 | 90,473 | | 8,142 | 8,262 | 9,192 | 10,060 | 10,798 | | 10,754 | 13,485 | 14,414 | 15,283 | 16,021 |
| | Integrated care - high | | 32,748 | 37,479 | 45,722 | 53,247 | 59,415 | | 8,142 | 5,426 | 6,036 | 6,607 | 7,091 | | 10,754 | 10,648 | 11,259 | 11,829 | 12,314 |
| 5 to 9 | Stable poverty projection | 30,258 | 35,783 | 43,196 | 49,745 | 56,769 | 62,772 | 3,870 | 4,147 | 4,558 | 4,937 | 5,327 | 5,676 | 8,635 | 8,913 | 9,324 | 9,702 | 10,093 | 10,442 |
| | 99% CI lower | 30,027 | 35,540 | 42,913 | 49,385 | 56,369 | 62,297 | 3,862 | 4,139 | 4,547 | 4,921 | 5,307 | 5,651 | 8,597 | 8,873 | 9,281 | 9,656 | 10,041 | 10,386 |
| | 99% CI upper | 30,488 | 36,026 | 43,479 | 50,105 | 57,170 | 63,247 | 3,888 | 4,168 | 4,582 | 4,964 | 5,359 | 5,711 | 8,684 | 8,965 | 9,379 | 9,761 | 10,156 | 10,508 |
| | % change - poverty stable | 0% | 18% | 43% | 64% | 88% | 107% | 0% | 7% | 18% | 28% | 38% | 47% | 0% | 3% | 8% | 12% | 17% | 21% |
| | Decreasing poverty projection | 29,982 | 35,380 | 42,282 | 48,336 | 54,875 | 60,424 | 3,892 | 4,015 | 4,358 | 4,743 | 5,137 | 5,493 | 8,658 | 8,780 | 9,123 | 9,508 | 9,903 | 10,258 |
| | 99%CI lower | 29,706 | 35,128 | 42,033 | 48,111 | 54,624 | 60,153 | 3,882 | 4,000 | 4,335 | 4,713 | 5,100 | 5,449 | 8,616 | 8,734 | 9,069 | 9,447 | 9,834 | 10,183 |
| | 99%CI upper | 30,096 | 35,504 | 42,430 | 48,528 | 55,107 | 60,682 | 3,914 | 4,032 | 4,372 | 4,765 | 5,167 | 5,529 | 8,711 | 8,828 | 9,169 | 9,562 | 9,964 | 10,326 |
| | % change - decreasing poverty | 0% | 18% | 41% | 61% | 83% | 102% | 0% | 3% | 12% | 22% | 32% | 41% | 0% | 1% | 5% | 10% | 14% | 18% |
| | Increasing poverty projection | 30,473 | 36,038 | 44,089 | 51,320 | 59,076 | 65,704 | 3,875 | 4,293 | 4,726 | 5,099 | 5,484 | 5,827 | 8,640 | 9,059 | 9,492 | 9,864 | 10,250 | 10,593 |
| | 99%CI lower | 30,463 | 36,028 | 44,079 | 51,310 | 59,066 | 65,694 | 3,874 | 4,291 | 4,721 | 5,091 | 5,475 | 5,816 | 8,608 | 9,025 | 9,455 | 9,826 | 10,209 | 10,550 |
| | 99%CI upper | 30,483 | 36,048 | 44,099 | 51,330 | 59,087 | 65,715 | 3,889 | 4,306 | 4,736 | 5,106 | 5,493 | 5,838 | 8,686 | 9,103 | 9,533 | 9,903 | 10,290 | 10,635 |
| | % change - increasing poverty | 0% | 18% | 45% | 68% | 94% | 116% | 0% | 11% | 22% | 32% | 42% | 50% | 0% | 5% | 10% | 14% | 19% | 23% |
| | COVID shock | | 17,892 | 43,196 | 49,745 | 56,769 | 62,772 | | 3,111 | 4,558 | 4,937 | 5,327 | 5,676 | | 5,493 | 9,324 | 9,702 | 10,093 | 10,442 |
| | Integrated care - moderate | | 17,892 | 34,341 | 39,547 | 45,131 | 49,904 | | 3,111 | 3,624 | 3,925 | 4,235 | 4,513 | | 5,493 | 8,389 | 8,690 | 9,001 | 9,278 |
| | Integrated care - high | | 17,892 | 25,485 | 29,349 | 33,494 | 37,036 | | 3,111 | 2,689 | 2,913 | 3,143 | 3,349 | | 5,493 | 7,455 | 7,678 | 7,909 | 8,114 |
| 10 to 14 | Stable poverty projection | 34,471 | 35,557 | 39,468 | 43,781 | 47,454 | 51,518 | 3,732 | 3,898 | 4,195 | 4,497 | 4,774 | 5,061 | 7,989 | 8,155 | 8,452 | 8,754 | 9,031 | 9,318 |
| | 99% CI lower | 34,322 | 35,010 | 38,686 | 42,842 | 46,302 | 50,210 | 3,690 | 3,838 | 4,123 | 4,389 | 4,636 | 4,884 | 7,916 | 8,063 | 8,348 | 8,615 | 8,861 | 9,109 |
| | 99% CI upper | 34,623 | 36,208 | 40,253 | 44,539 | 48,324 | 52,376 | 3,757 | 3,959 | 4,281 | 4,600 | 4,899 | 5,203 | 8,046 | 8,248 | 8,570 | 8,889 | 9,188 | 9,491 |
| | % change - poverty stable | 0% | 3% | 14% | 27% | 38% | 49% | 0% | 4% | 12% | 21% | 28% | 36% | 0% | 2% | 6% | 10% | 13% | 17% |
| | Decreasing poverty projection | 34,527 | 33,345 | 37,064 | 41,539 | 45,549 | 49,786 | 3,735 | 3,736 | 3,968 | 4,275 | 4,558 | 4,849 | 7,992 | 7,993 | 8,225 | 8,532 | 8,815 | 9,106 |
| | 99%CI lower | 34,045 | 33,199 | 36,829 | 41,213 | 45,139 | 49,289 | 3,726 | 3,530 | 3,684 | 3,991 | 4,266 | 4,556 | 7,951 | 7,756 | 7,909 | 8,216 | 8,491 | 8,782 |
| | 99%CI upper | 34,639 | 33,394 | 37,136 | 41,635 | 45,667 | 49,927 | 3,745 | 3,942 | 4,253 | 4,561 | 4,850 | 5,143 | 8,033 | 8,231 | 8,542 | 8,849 | 9,139 | 9,431 |
| | % change - decreasing poverty | 0% | -3% | 7% | 20% | 32% | 44% | 0% | 0% | 6% | 14% | 22% | 30% | 0% | 0% | 3% | 7% | 10% | 14% |
| | Increasing poverty projection | 34,697 | 39,666 | 45,331 | 49,763 | 53,615 | 57,800 | 3,727 | 4,120 | 4,449 | 4,751 | 5,011 | 5,295 | 7,984 | 8,377 | 8,706 | 9,008 | 9,268 | 9,552 |
| | 99%CI lower | 34,593 | 39,113 | 44,632 | 48,902 | 52,600 | 56,627 | 3,726 | 4,119 | 4,448 | 4,750 | 5,009 | 5,294 | 7,951 | 8,345 | 8,673 | 8,975 | 9,235 | 9,520 |
| | 99%CI upper | 34,736 | 39,795 | 45,496 | 49,965 | 53,851 | 58,070 | 3,728 | 4,121 | 4,450 | 4,752 | 5,012 | 5,296 | 8,016 | 8,410 | 8,739 | 9,040 | 9,300 | 9,585 |
| | % change - increasing poverty | 0% | 14% | 31% | 43% | 55% | 67% | 0% | 11% | 19% | 27% | 34% | 42% | 0% | 5% | 9% | 13% | 16% | 20% |
| | COVID shock | | 17,778 | 39,468 | 43,781 | 47,454 | 51,518 | | 2,923 | 4,195 | 4,497 | 4,774 | 5,061 | | 5,052 | 8,452 | 8,754 | 9,031 | 9,318 |
| | Integrated care - moderate | | 17,778 | 34,930 | 38,746 | 41,997 | 45,594 | | 2,923 | 3,713 | 3,980 | 4,225 | 4,479 | | 5,052 | 7,970 | 8,237 | 8,482 | 8,736 |
| | Integrated care - high | | 17,778 | 30,391 | 33,711 | 36,540 | 39,669 | | 2,923 | 3,230 | 3,463 | 3,676 | 3,897 | | 5,052 | 7,487 | 7,720 | 7,933 | 8,154 |
| 15 to 19 | Stable poverty projection | 38,014 | 42,972 | 63,316 | 73,247 | 81,796 | 91,061 | 5,138 | 5,234 | 5,248 | 5,356 | 5,479 | 5,581 | 10,132 | 10,228 | 10,242 | 10,351 | 10,473 | 10,575 |
| | 99% CI lower | 37,675 | 42,341 | 62,358 | 71,889 | 80,024 | 88,915 | 5,035 | 5,108 | 5,230 | 5,336 | 5,442 | 5,542 | 9,996 | 10,069 | 10,191 | 10,297 | 10,403 | 10,503 |
| | 99% CI upper | 38,462 | 43,763 | 64,485 | 74,882 | 83,915 | 93,620 | 5,226 | 5,341 | 5,305 | 5,437 | 5,571 | 5,693 | 10,253 | 10,368 | 10,333 | 10,464 | 10,598 | 10,721 |
| | % change - poverty stable | 0% | 13% | 67% | 93% | 115% | 140% | 0% | 2% | 2% | 4% | 7% | 9% | 0% | 1% | 1% | 2% | 3% | 4% |
| | Decreasing poverty projection | 37,726 | 40,829 | 61,093 | 70,520 | 78,481 | 87,276 | 5,217 | 4,960 | 4,792 | 4,921 | 5,069 | 5,190 | 10,211 | 9,954 | 9,786 | 9,915 | 10,063 | 10,184 |
| | 99%CI lower | 37,464 | 40,719 | 60,930 | 70,308 | 78,220 | 86,975 | 5,169 | 4,902 | 4,703 | 4,813 | 4,940 | 5,005 | 10,130 | 9,863 | 9,664 | 9,774 | 9,900 | 9,966 |
| | 99%CI upper | 37,978 | 41,070 | 61,485 | 71,050 | 79,142 | 88,067 | 5,296 | 5,040 | 4,872 | 5,044 | 5,240 | 5,366 | 10,323 | 10,068 | 9,900 | 10,072 | 10,267 | 10,394 |
| | % change - decreasing poverty | 0% | 8% | 62% | 87% | 108% | 131% | 0% | -5% | -8% | -6% | -3% | -1% | 0% | -3% | -4% | -3% | -1% | 0% |
| | Increasing poverty projection | 37,197 | 43,225 | 64,357 | 73,790 | 82,117 | 91,299 | 5,166 | 5,471 | 5,496 | 5,615 | 5,753 | 5,865 | 10,160 | 10,466 | 10,490 | 10,609 | 10,747 | 10,859 |
| | 99%CI lower | 37,121 | 42,800 | 63,015 | 72,146 | 80,268 | 89,190 | 5,106 | 5,210 | 5,331 | 5,472 | 5,617 | 5,749 | 10,067 | 10,171 | 10,292 | 10,433 | 10,578 | 10,710 |
| | 99%CI upper | 37,352 | 43,805 | 65,789 | 75,329 | 83,705 | 92,979 | 5,225 | 5,731 | 5,660 | 5,757 | 5,888 | 5,979 | 10,253 | 10,759 | 10,688 | 10,784 | 10,915 | 11,006 |
| | % change - increasing poverty | 0% | 16% | 73% | 98% | 121% | 145% | 0% | 6% | 6% | 9% | 11% | 14% | 0% | 3% | 3% | 4% | 6% | 7% |
| | COVID shock | | 21,486 | 63,316 | 73,247 | 81,796 | 91,061 | | 3,925 | 5,248 | 5,356 | 5,479 | 5,581 | | 6,422 | 10,242 | 10,351 | 10,473 | 10,575 |
| | Integrated care - moderate | | 21,486 | 57,618 | 66,655 | 74,434 | 82,866 | | 3,925 | 4,775 | 4,874 | 4,986 | 5,079 | | 6,422 | 9,769 | 9,869 | 9,980 | 10,073 |
| | Integrated care - high | | 21,486 | 51,919 | 60,063 | 67,073 | 74,670 | | 3,925 | 4,303 | 4,392 | 4,493 | 4,576 | | 6,422 | 9,297 | 9,386 | 9,487 | 9,571 |

**Appendix Table 7. Change in estimated total admissions and ED attendances in 2030 and 2040 for COVID integrated care scenarios compared with stable, decreasing and increasing poverty scenarios**

| Modelled scenario | COVID scenario | infant | | 1-4y | | 5-9y | | 10-14y | | 15-19y | |
|---|---|---|---|---|---|---|---|---|---|---|---|
| **Total admissions** | | **2030** | **2040** | **2030** | **2040** | **2030** | **2040** | **2030** | **2040** | **2030** | **2040** |
| Stable poverty | Integrated-moderate | -20.8% | -21.2% | -23.9% | -24.9% | -10.4% | -11.1% | -5.9% | -6.2% | -4.7% | -4.7% |
| | Integrated-high | -41.7% | -42.4% | -40.6% | -42.3% | -20.9% | -22.3% | -11.8% | -12.5% | -9.3% | -9.5% |
| Decreasing poverty | Integrated-moderate | -22.5% | -23.9% | -20.2% | -23.0% | -8.6% | -9.6% | -3.5% | -4.1% | -0.5% | -1.1% |
| | Integrated-high | -42.9% | -44.4% | -37.7% | -40.8% | -19.2% | -20.9% | -9.5% | -10.5% | -5.3% | -6.0% |
| Increaseing poverty | Integrated-moderate | -25.0% | -25.9% | -28.4% | -28.8% | -11.9% | -12.4% | -8.6% | -8.5% | -7.0% | -7.2% |
| | Integrated-high | -44.8% | -45.8% | -44.0% | -45.3% | -22.2% | -23.4% | -14.3% | -14.6% | -11.5% | -11.9% |
| **ED attendances** | | | | | | | | | | | |
| Stable poverty | Integrated-moderate | -23.5% | -23.5% | -33.0% | -33.0% | -20.5% | -20.5% | -11.5% | -11.5% | -9.0% | -9.0% |
| | Integrated-high | -47.0% | -47.0% | -56.0% | -56.0% | -41.0% | -41.0% | -23.0% | -23.0% | -18.0% | -18.0% |
| Decreasing poverty | Integrated-moderate | -29.9% | -30.1% | -30.0% | -29.8% | -18.2% | -17.4% | -6.7% | -8.4% | -5.5% | -5.1% |
| | Integrated-high | -51.4% | -51.6% | -54.0% | -53.9% | -39.3% | -38.7% | -18.8% | -20.3% | -14.8% | -14.4% |
| Increaseing poverty | Integrated-moderate | -25.5% | -26.2% | -31.1% | -30.8% | -22.9% | -24.0% | -22.1% | -21.1% | -9.7% | -9.2% |
| | Integrated-high | -48.4% | -48.9% | -54.7% | -54.6% | -42.8% | -43.6% | -32.3% | -31.4% | -18.6% | -18.2% |

#### Appendix Table 8. Outpatient attendance projections to 2040 in different poverty scenarios

| | | 2016 | 2017 | 2020 | 2025 | 2030 | 2035 | 2040 |
|---|---|---|---|---|---|---|---|---|
| infant | Recorded attendance rate | 140,183 | 138,690 | | | | | |
| | Stable poverty projection | 142,198 | 150,652 | 161,192 | 190,882 | 220,005 | 244,827 | 268,281 |
| | (stable lower bound) | 137,868 | 146,450 | 154,261 | 179,164 | 203,636 | 224,247 | 243,697 |
| | (stable higher bound) | 150,406 | 156,915 | 169,533 | 203,179 | 236,264 | 264,598 | 291,461 |
| | % rise stable poverty scenario | 0% | 6% | 13% | 34% | 55% | 72% | 89% |
| | Decreasing poverty projection | 141,115 | 151,574 | 162,575 | 193,408 | 223,756 | 249,699 | 274,224 |
| | (decreasing lower bound) | 135,448 | 145,970 | 155,650 | 184,930 | 214,848 | 240,586 | 264,063 |
| | (decreasing higher bound) | 148,398 | 159,383 | 171,902 | 205,612 | 239,004 | 268,487 | 295,450 |
| | % rise decreasing poverty scenario | 0% | 7% | 15% | 37% | 59% | 77% | 94% |
| | Increasing poverty projection | 141,115 | 151,574 | 163,076 | 194,065 | 224,404 | 250,330 | 274,839 |
| | (increasing lower bound) | 135,448 | 145,970 | 158,447 | 189,414 | 218,497 | 243,283 | 266,558 |
| | (increasing upper bound) | 148,398 | 159,383 | 173,824 | 207,692 | 240,684 | 269,737 | 297,022 |
| | % rise increasing poverty scenario | 0% | 7% | 16% | 38% | 59% | 77% | 95% |
| 1 to 4 | Recorded attendance rate | 75,616 | 73,322 | | | | | |
| | Stable poverty projection | 75,494 | 73,268 | 74,037 | 79,003 | 81,918 | 86,324 | 91,253 |
| | (stable lower bound) | 75,104 | 72,724 | 73,166 | 77,599 | 79,986 | 83,899 | 88,364 |
| | (stable higher bound) | 75,869 | 73,855 | 75,117 | 80,607 | 84,214 | 89,136 | 94,468 |
| | % rise stable poverty scenario | 0% | -3% | -2% | 5% | 9% | 14% | 21% |
| | Decreasing poverty projection | 75,494 | 73,268 | 65,789 | 67,234 | 70,247 | 74,899 | 80,133 |
| | (decreasing lower bound) | 75,104 | 72,724 | 64,618 | 65,403 | 67,892 | 72,060 | 76,840 |
| | (decreasing higher bound) | 75,869 | 73,855 | 67,289 | 69,452 | 73,133 | 78,289 | 83,910 |
| | % rise decreasing poverty scenario | 0% | -3% | -13% | -11% | -7% | -1% | 6% |
| | Increasing poverty projection | 75,494 | 73,268 | 81,650 | 87,799 | 90,640 | 94,863 | 99,565 |
| | (increasing lower bound) | 75,104 | 72,724 | 80,549 | 86,426 | 88,901 | 92,744 | 96,977 |
| | (increasing upper bound) | 75,869 | 73,855 | 82,420 | 89,095 | 92,637 | 97,302 | 102,386 |
| | % rise increasing poverty scenario | 0% | -3% | 8% | 16% | 20% | 26% | 32% |
| 5 to 9 | Recorded attendance rate | 71,256 | 68,535 | | | | | |
| | Stable poverty projection | 69,673 | 71,116 | 81,578 | 95,451 | 107,779 | 120,930 | 132,238 |
| | (stable lower bound) | 68,359 | 69,542 | 78,467 | 90,186 | 100,564 | 111,671 | 121,187 |
| | (stable higher bound) | 70,923 | 72,585 | 83,732 | 98,818 | 112,319 | 126,627 | 139,022 |
| | % rise stable poverty scenario | 0% | 2% | 17% | 37% | 55% | 74% | 90% |
| | Decreasing poverty projection | 69,673 | 71,116 | 80,286 | 93,579 | 105,947 | 119,114 | 130,462 |
| | (decreasing lower bound) | 68,355 | 69,538 | 77,457 | 88,725 | 99,134 | 110,252 | 119,798 |
| | (decreasing higher bound) | 70,929 | 72,591 | 82,093 | 96,441 | 109,993 | 124,325 | 136,772 |
| | % rise decreasing poverty scenario | 0% | 2% | 15% | 34% | 52% | 71% | 87% |
| | Increasing poverty projection | 69,673 | 71,116 | 82,770 | 96,850 | 109,149 | 122,287 | 133,566 |
| | (increasing lower bound) | 68,355 | 69,538 | 79,390 | 91,268 | 101,622 | 112,715 | 122,208 |
| | (increasing upper bound) | 70,929 | 72,591 | 85,256 | 100,608 | 114,073 | 128,453 | 140,828 |
| | % rise increasing poverty scenario | 0% | 2% | 19% | 39% | 57% | 76% | 92% |
| 10 to 14 | Recorded attendance rate | 76,993 | 74,774 | | | | | |
| | Stable poverty projection | 75,231 | 76,331 | 80,573 | 90,084 | 100,069 | 108,960 | 118,409 |
| | (stable lower bound) | 74,395 | 75,560 | 79,121 | 88,216 | 98,015 | 106,538 | 115,790 |
| | (stable higher bound) | 75,557 | 77,097 | 82,499 | 92,618 | 102,968 | 112,413 | 122,236 |
| | % rise stable poverty scenario | 0% | 1% | 7% | 20% | 33% | 45% | 57% |
| | Decreasing poverty projection | 75,231 | 76,331 | 79,820 | 89,009 | 99,002 | 107,916 | 117,374 |
| | (decreasing lower bound) | 74,138 | 75,385 | 76,873 | 85,012 | 94,835 | 103,423 | 112,705 |
| | (decreasing higher bound) | 75,696 | 77,153 | 82,518 | 92,639 | 102,986 | 112,430 | 122,260 |
| | % rise decreasing poverty scenario | 0% | 1% | 6% | 18% | 32% | 43% | 56% |
| | Increasing poverty projection | 75,231 | 76,331 | 81,268 | 90,887 | 100,866 | 109,741 | 119,183 |
| | (increasing lower bound) | 74,138 | 75,385 | 79,463 | 88,660 | 98,217 | 106,730 | 115,807 |
| | (increasing upper bound) | 75,696 | 77,153 | 82,575 | 92,747 | 103,032 | 112,475 | 122,303 |
| | % rise increasing poverty scenario | 0% | 1% | 8% | 21% | 34% | 46% | 58% |
| 15 to 19 | Recorded attendance rate | 77,714 | 77,177 | | | | | |
| | Stable poverty projection | 77,582 | 78,258 | 89,165 | 120,382 | 139,253 | 156,534 | 174,147 |
| | (stable lower bound) | 76,520 | 77,298 | 87,633 | 117,335 | 135,197 | 151,537 | 168,207 |
| | (stable higher bound) | 78,400 | 79,166 | 90,630 | 122,702 | 142,373 | 160,453 | 178,812 |
| | % rise stable poverty scenario | 0% | 1% | 15% | 55% | 79% | 102% | 124% |
| | Decreasing poverty projection | 76,569 | 77,017 | 84,273 | 114,838 | 133,245 | 149,887 | 167,065 |
| | (decreasing lower bound) | 76,087 | 76,295 | 81,852 | 111,040 | 128,653 | 144,504 | 160,941 |
| | (decreasing higher bound) | 77,352 | 77,806 | 86,959 | 119,560 | 138,461 | 155,548 | 173,196 |
| | % rise decreasing poverty scenario | 0% | 1% | 10% | 50% | 74% | 96% | 118% |
| | Increasing poverty projection | 76,569 | 77,017 | 90,231 | 121,877 | 140,137 | 156,732 | 173,774 |
| | (increasing lower bound) | 76,087 | 76,295 | 88,031 | 120,706 | 138,118 | 153,904 | 170,155 |
| | (increasing upper bound) | 77,352 | 77,806 | 92,426 | 123,900 | 142,428 | 159,381 | 176,675 |
| | % rise increasing poverty scenario | 0% | 1% | 18% | 59% | 83% | 105% | 127% |

### Appendix Table 9. Inputs for outpatient projection models

| | | Year of year | | | | | | | | | | | | | | | | | | | |
|---|---|---|---|---|---|---|---|---|---|---|---|---|---|---|---|---|---|---|---|---|---|
| | | 2007 | 2008 | 2009 | 2010 | 2011 | 2012 | 2013 | 2014 | 2015 | 2016 | 2017 | 2018 | 2019 | 2020 | 2021 | 2022 | 2025 | 2030 | 2035 | 2040 |
| infant | Number of attendances | 517,618 | 584,585 | 627,514 | 649,754 | 696,137 | 729,757 | 766,342 | 875,492 | 997,941 | 937,967 | 906,296 | | | | | | | | | |
| | Population | 642,603 | 667,840 | 662,806 | 672,051 | 679,102 | 696,441 | 676,531 | 664,183 | 662,977 | 669,103 | 653,467 | 686,611 | 690,746 | 694,041 | 696,769 | 699,510 | 703,340 | 711,958 | 731,170 | 750,673 |
| | Recorded attendance rate | 80,550 | 87,534 | 94,675 | 96,682 | 102,508 | 104,784 | 113,275 | 131,815 | 150,524 | 140,183 | 138,690 | | | | | | | | | |
| | Child poverty rate | 32.0% | 31.0% | 30.0% | 27.5% | 27.5% | 27.8% | 27.5% | 28.0% | 29.0% | 30.5% | 29.5% | | | | | | | | | |
| | Poverty: stable poverty scenario | 32.0% | 31.0% | 30.0% | 27.5% | 27.5% | 27.8% | 27.5% | 28.0% | 29.0% | 30.5% | 29.5% | 29.5% | 29.5% | 29.5% | 29.5% | 29.5% | 29.5% | 29.5% | 29.5% | 29.5% |
| | Poverty: decreasing poverty scenario | 32.0% | 31.0% | 30.0% | 27.5% | 27.5% | 27.8% | 27.5% | 28.0% | 29.0% | 30.5% | 29.5% | 27.0% | 25.0% | 23.0% | 20.0% | 20.0% | 20.0% | 20.0% | 20.0% | 20.0% |
| | Poverty: increasing poverty scenario | 32.0% | 31.0% | 30.0% | 27.5% | 27.5% | 27.8% | 27.5% | 28.0% | 29.0% | 30.5% | 29.5% | 33.0% | 34.5% | 35.5% | 36.6% | 36.6% | 36.6% | 36.6% | 36.6% | 36.6% |
| 1 to 4 | Number of attendances | 1,316,405 | 1,442,806 | 1,537,095 | 1,617,019 | 1,705,739 | 1,789,734 | 1,891,170 | 1,999,017 | 2,073,714 | 2,086,964 | 2,002,757 | | | | | | | | | |
| | Population | 2,406,417 | 2,474,865 | 2,549,099 | 2,608,443 | 2,649,644 | 2,696,915 | 2,737,599 | 2,766,774 | 2,771,703 | 2,759,943 | 2,731,458 | 2,681,826 | 2,702,026 | 2,723,418 | 2,742,475 | 2,757,687 | 2,789,355 | 2,812,851 | 2,873,331 | 2,952,022 |
| | Recorded attendance rate | 54,704 | 58,298 | 60,300 | 61,992 | 64,376 | 66,362 | 69,081 | 72,251 | 74,817 | 75,616 | 73,322 | | | | | | | | | |
| | Child poverty rate | 32.0% | 31.0% | 30.0% | 27.5% | 27.5% | 27.8% | 27.5% | 28.0% | 29.0% | 30.5% | 29.5% | | | | | | | | | |
| | Poverty: stable poverty scenario | 32.0% | 31.0% | 30.0% | 27.5% | 27.5% | 27.8% | 27.5% | 28.0% | 29.0% | 30.5% | 29.5% | 29.5% | 29.5% | 29.5% | 29.5% | 29.5% | 29.5% | 29.5% | 29.5% | 29.5% |
| | Poverty: decreasing poverty scenario | 32.0% | 31.0% | 30.0% | 27.5% | 27.5% | 27.8% | 27.5% | 28.0% | 29.0% | 30.5% | 29.5% | 27.0% | 25.0% | 23.0% | 20.0% | 20.0% | 20.0% | 20.0% | 20.0% | 20.0% |
| | Poverty: increasing poverty scenario | 32.0% | 31.0% | 30.0% | 27.5% | 27.5% | 27.8% | 27.5% | 28.0% | 29.0% | 30.5% | 29.5% | 33.0% | 34.5% | 35.5% | 36.6% | 36.6% | 36.6% | 36.6% | 36.6% | 36.6% |
| 5 to 9 | Number of attendances | 1,460,220 | 1,549,755 | 1,649,705 | 1,734,872 | 1,823,304 | 1,917,380 | 2,068,698 | 2,192,522 | 2,288,576 | 2,442,849 | 2,396,948 | | | | | | | | | |
| | Population | 2,937,497 | 2,912,054 | 2,911,772 | 2,934,351 | 2,990,135 | 3,083,582 | 3,187,919 | 3,272,365 | 3,357,463 | 3,428,266 | 3,497,402 | 3,416,423 | 3,414,701 | 3,405,025 | 3,392,541 | 3,381,758 | 3,433,837 | 3,508,479 | 3,540,518 | 3,618,967 |
| | Recorded attendance rate | 49,710 | 53,219 | 56,656 | 59,123 | 60,977 | 62,180 | 64,892 | 67,001 | 68,164 | 71,256 | 68,535 | | | | | | | | | |
| | Child poverty rate | 32.0% | 31.0% | 30.0% | 27.5% | 27.5% | 27.8% | 27.5% | 28.0% | 29.0% | 30.5% | 29.5% | | | | | | | | | |
| | Poverty: stable poverty scenario | 32.0% | 31.0% | 30.0% | 27.5% | 27.5% | 27.8% | 27.5% | 28.0% | 29.0% | 30.5% | 29.5% | 29.5% | 29.5% | 29.5% | 29.5% | 29.5% | 29.5% | 29.5% | 29.5% | 29.5% |
| | Poverty: decreasing poverty scenario | 32.0% | 31.0% | 30.0% | 27.5% | 27.5% | 27.8% | 27.5% | 28.0% | 29.0% | 30.5% | 29.5% | 27.0% | 25.0% | 23.0% | 20.0% | 20.0% | 20.0% | 20.0% | 20.0% | 20.0% |
| | Poverty: increasing poverty scenario | 32.0% | 31.0% | 30.0% | 27.5% | 27.5% | 27.8% | 27.5% | 28.0% | 29.0% | 30.5% | 29.5% | 33.0% | 34.5% | 35.5% | 36.6% | 36.6% | 36.6% | 36.6% | 36.6% | 36.6% |
| 10 to 14 | Number of attendances | 1,690,843 | 1,774,005 | 1,852,712 | 1,894,701 | 1,936,944 | 1,943,652 | 2,038,663 | 2,123,202 | 2,212,887 | 2,363,889 | 2,367,388 | | | | | | | | | |
| | Population | 3,167,468 | 3,150,742 | 3,128,839 | 3,109,239 | 3,067,411 | 3,007,871 | 2,976,393 | 2,973,055 | 3,000,295 | 3,070,254 | 3,166,038 | 3,226,138 | 3,298,733 | 3,369,859 | 3,420,039 | 3,459,723 | 3,449,793 | 3,476,225 | 3,550,753 | 3,582,470 |
| | Recorded attendance rate | 53,382 | 56,304 | 59,214 | 60,938 | 63,146 | 64,619 | 68,494 | 71,415 | 73,756 | 76,993 | 74,774 | | | | | | | | | |
| | Child poverty rate | 32.0% | 31.0% | 30.0% | 27.5% | 27.5% | 27.8% | 27.5% | 28.0% | 29.0% | 30.5% | 29.5% | | | | | | | | | |
| | Poverty: stable poverty scenario | 32.0% | 31.0% | 30.0% | 27.5% | 27.5% | 27.8% | 27.5% | 28.0% | 29.0% | 30.5% | 29.5% | 29.5% | 29.5% | 29.5% | 29.5% | 29.5% | 29.5% | 29.5% | 29.5% | 29.5% |
| | Poverty: decreasing poverty scenario | 32.0% | 31.0% | 30.0% | 27.5% | 27.5% | 27.8% | 27.5% | 28.0% | 29.0% | 30.5% | 29.5% | 27.0% | 25.0% | 23.0% | 20.0% | 20.0% | 20.0% | 20.0% | 20.0% | 20.0% |
| | Poverty: increasing poverty scenario | 32.0% | 31.0% | 30.0% | 27.5% | 27.5% | 27.8% | 27.5% | 28.0% | 29.0% | 30.5% | 29.5% | 33.0% | 34.5% | 35.5% | 36.6% | 36.6% | 36.6% | 36.6% | 36.6% | 36.6% |
| 15 to 19 | Number of attendances | 1,749,617 | 1,885,345 | 2,026,484 | 2,053,075 | 2,087,413 | 2,129,108 | 2,257,488 | 2,315,286 | 2,356,004 | 2,470,860 | 2,408,498 | | | | | | | | | |
| | Population | 3,332,566 | 3,337,169 | 3,355,015 | 3,343,131 | 3,324,270 | 3,286,306 | 3,254,752 | 3,230,954 | 3,213,289 | 3,179,410 | 3,120,730 | 3,100,022 | 3,088,720 | 3,104,613 | 3,163,138 | 3,248,912 | 3,489,328 | 3,563,706 | 3,588,090 | 3,661,150 |
| | Recorded attendance rate | 52,501 | 56,495 | 60,402 | 61,412 | 62,793 | 64,787 | 69,360 | 71,660 | 73,321 | 77,714 | 77,177 | | | | | | | | | |
| | Child poverty rate | 32.0% | 31.0% | 30.0% | 27.5% | 27.5% | 27.8% | 27.5% | 28.0% | 29.0% | 30.5% | 29.5% | | | | | | | | | |
| | Poverty: stable poverty scenario | 32.0% | 31.0% | 30.0% | 27.5% | 27.5% | 27.8% | 27.5% | 28.0% | 29.0% | 30.5% | 29.5% | 29.5% | 29.5% | 29.5% | 29.5% | 29.5% | 29.5% | 29.5% | 29.5% | 29.5% |
| | Poverty: decreasing poverty scenario | 32.0% | 31.0% | 30.0% | 27.5% | 27.5% | 27.8% | 27.5% | 28.0% | 29.0% | 30.5% | 29.5% | 27.0% | 25.0% | 23.0% | 20.0% | 20.0% | 20.0% | 20.0% | 20.0% | 20.0% |
| | Poverty: increasing poverty scenario | 32.0% | 31.0% | 30.0% | 27.5% | 27.5% | 27.8% | 27.5% | 28.0% | 29.0% | 30.5% | 29.5% | 33.0% | 34.5% | 35.5% | 36.6% | 36.6% | 36.6% | 36.6% | 36.6% | 36.6% |

**Appendix Table 10. Model parameters for outpatient projection models**

| | | Mean | Std. Dev. | Median | [95% Credible interval] | |
|---|---|---|---|---|---|---|
| Infant | Year | 44758.71 | 4615.201 | 42919.12 | 37948.26 | 50784.24 |
| | Child poverty | 1175081 | 1681565 | 430231 | 0.0003101 | 3274893 |
| | intercept | 190226.1 | 513552 | 423629.1 | -455240.5 | 539999.1 |
| 1-4 years | Year | 10497.94 | 2553.45 | 10339.31 | 7523.589 | 14489.74 |
| | Population | 2.045105 | 0.0503626 | 2.037918 | 1.961953 | 2.107205 |
| | Child poverty | 3455668 | 152954 | 3522604 | 3235120 | 3581310 |
| | intercept | -4709231 | 147129.9 | -4711596 | -4852296 | -4453175 |
| 5-9 years | Year | 99480.13 | 13252.2 | 106502.6 | 84193.5 | 108877.2 |
| | Population | 0.0855366 | 0.074886 | 0.1162133 | 1.22E-07 | 0.1400925 |
| | Child poverty | 676637.2 | 170135.4 | 610996.4 | 526259.3 | 867566.3 |
| | intercept | 993649.6 | 216457.3 | 945286.1 | 800474.3 | 1244829 |
| 10-14 years | Year | 72037.44 | 1495.483 | 71499.13 | 69651.89 | 73960.62 |
| | Population | 0.4044908 | 0.1653421 | 0.3947823 | 0.2443095 | 0.5801009 |
| | Child poverty | 390427.7 | 666076.4 | 3639.534 | 9.568 | 1159372 |
| | intercept | 300453.2 | 367201.3 | 446649.8 | -134187.2 | 571482.3 |
| 15-19 years | Year | 115022.8 | 4954.09 | 117814.1 | 109274.6 | 118104.2 |
| | Population | 2.235418 | 0.0682365 | 2.204117 | 2.188326 | 2.317539 |
| | Child poverty | 1479729 | 964824.2 | 2030726 | 312299.6 | 2042550 |
| | intercept | -6159396 | 47125.03 | -6183998 | -6189126 | -6102577 |

#### Appendix Figure 1. ED attendance and emergency admission rates by region for 5-24 year olds

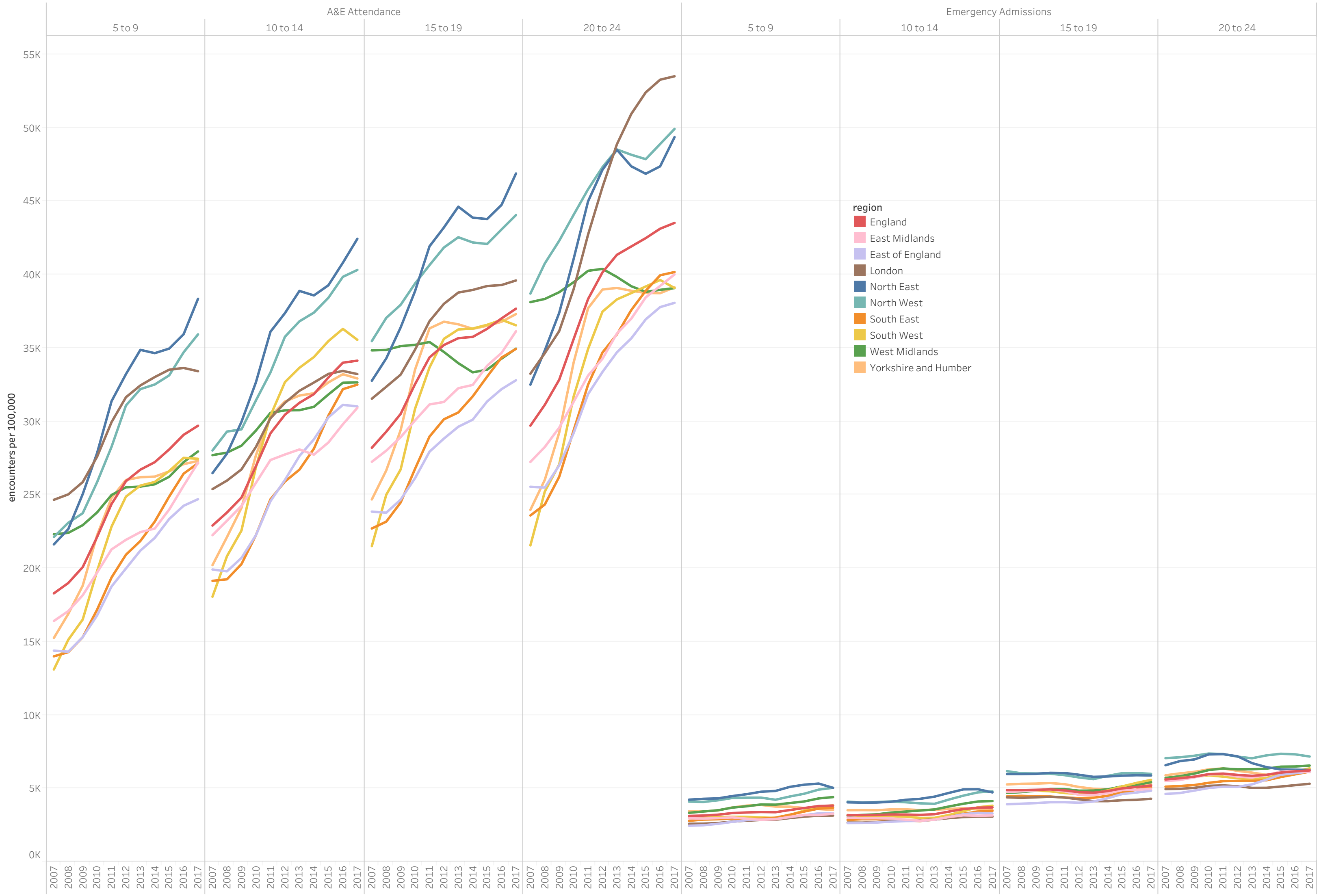

**Appendix Figure 2. Attendances and admission rates by deprivation quintile and age**

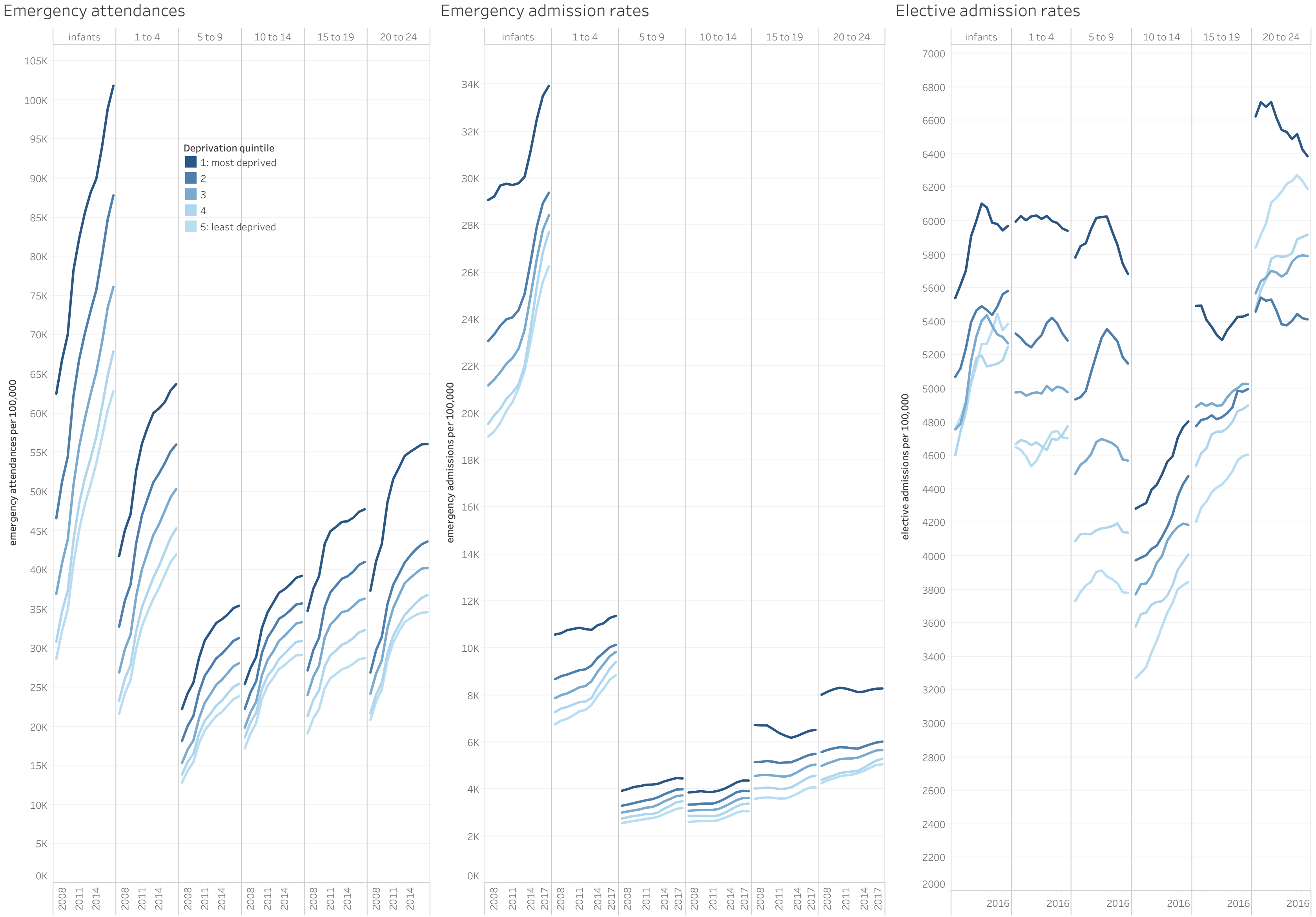

#### Appendix Figure 3. Emergency admissions by case, 5-14 years

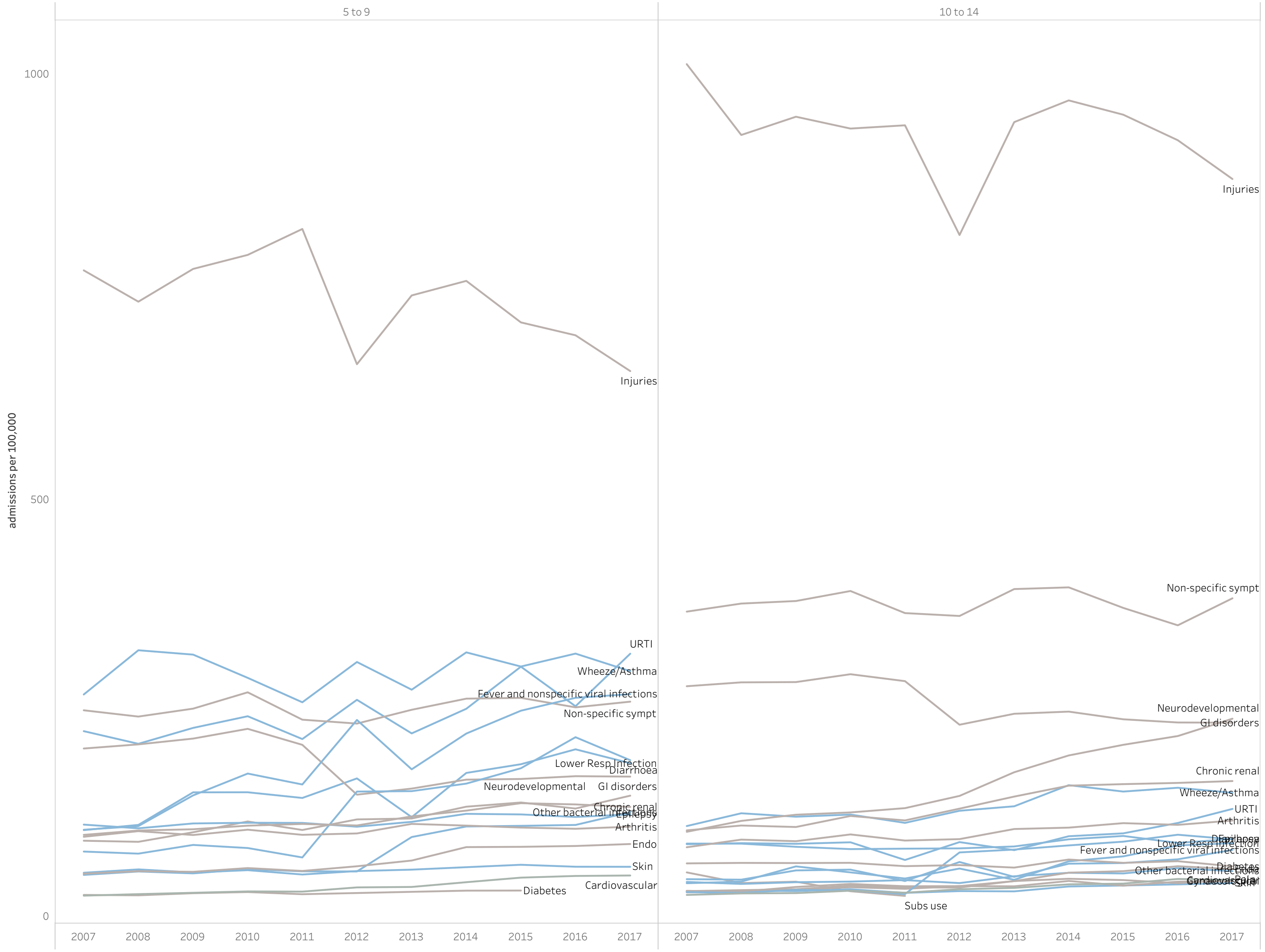

**Appendix Figure 4. Emergency admissions by cause, 15-24 years**

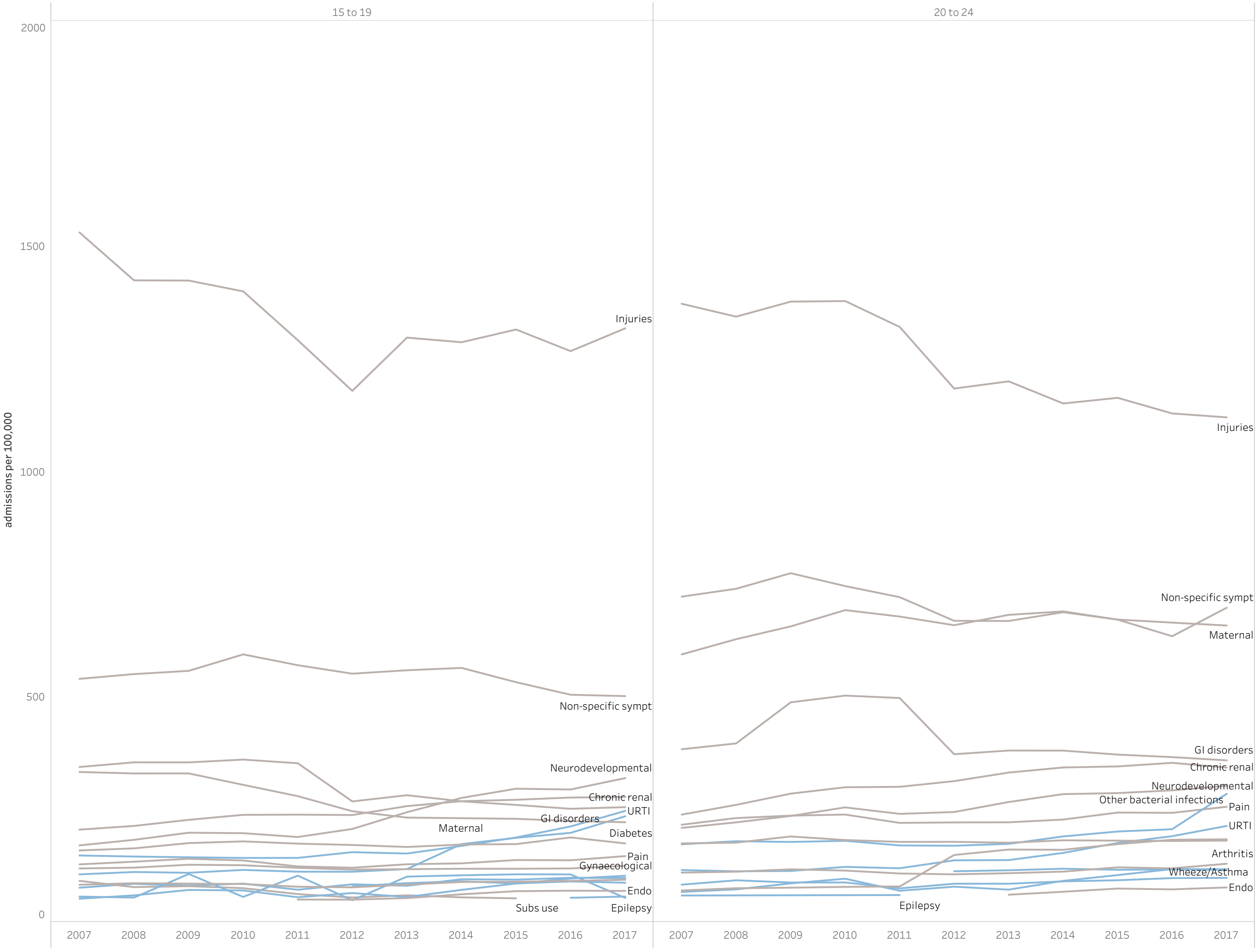

**Appendix Figure 5. Proportion of emergency admissions that are ACSC by age-group in England, 2007 to 2017**

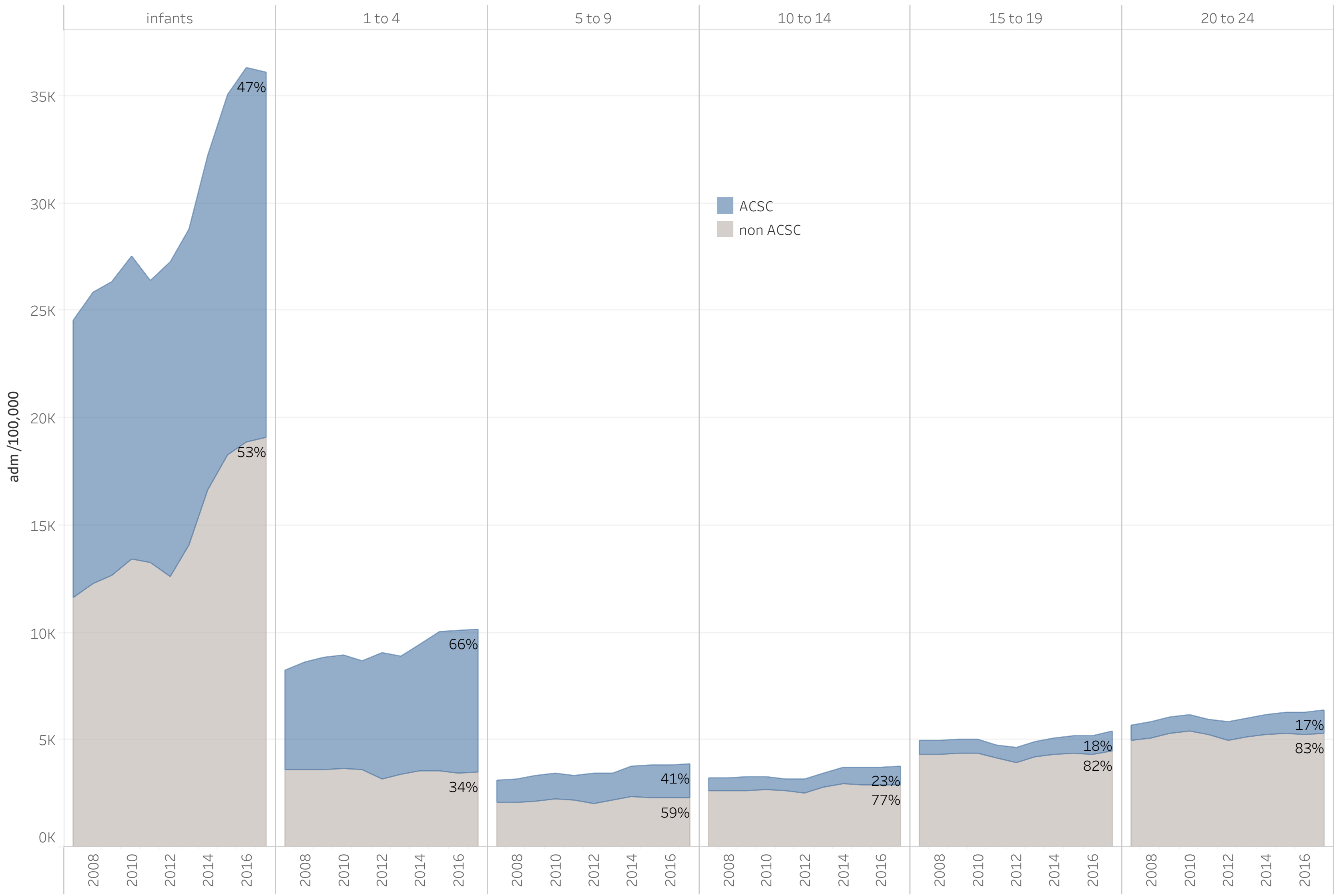

**Appendix Figure 6. Proportion of emergency admissions amongst infants that are ACSC by region**

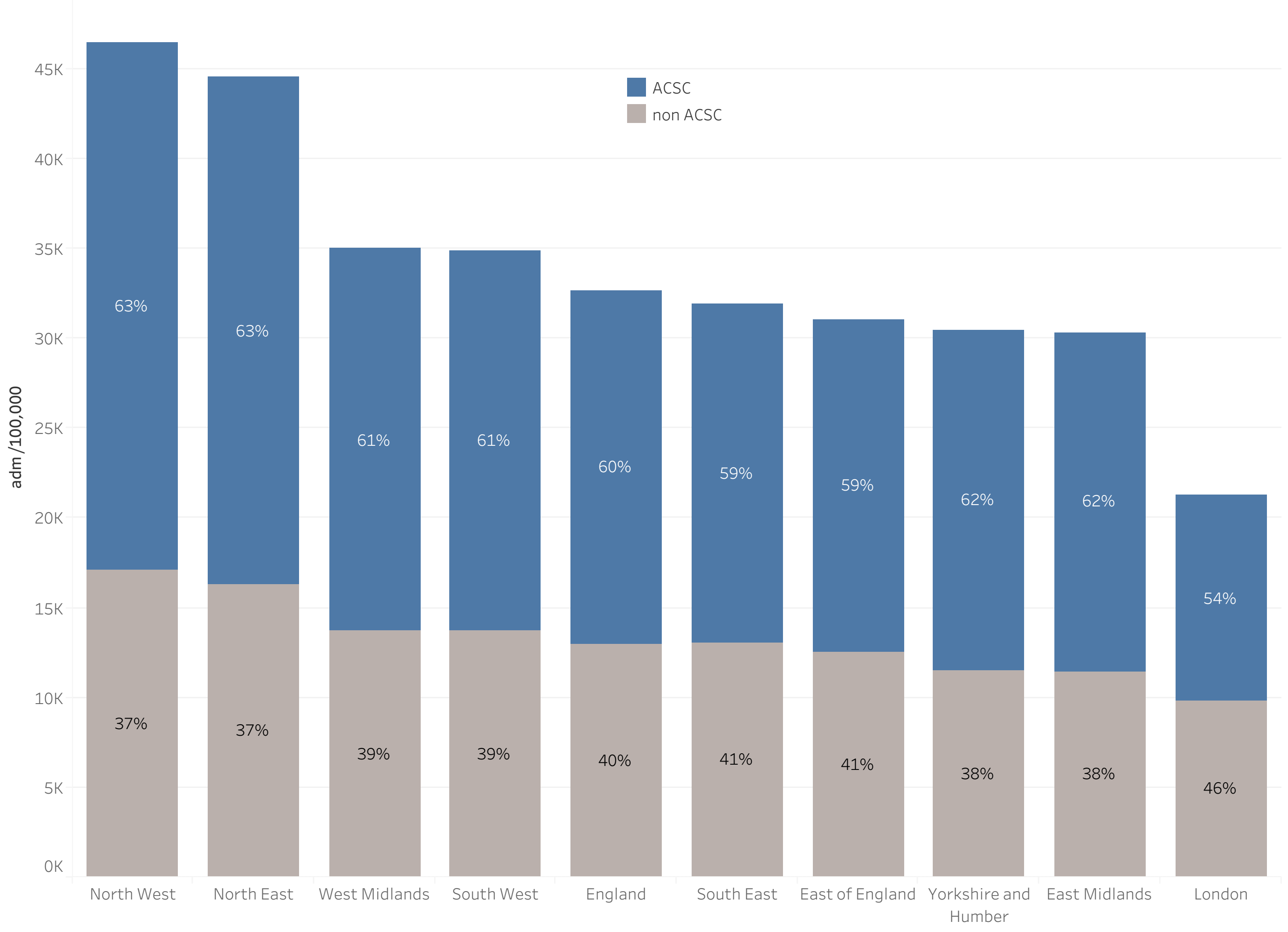

**Appendix Figure 7. Causes of elective admissions amongst 0-14 year olds, 2007 and 2017**

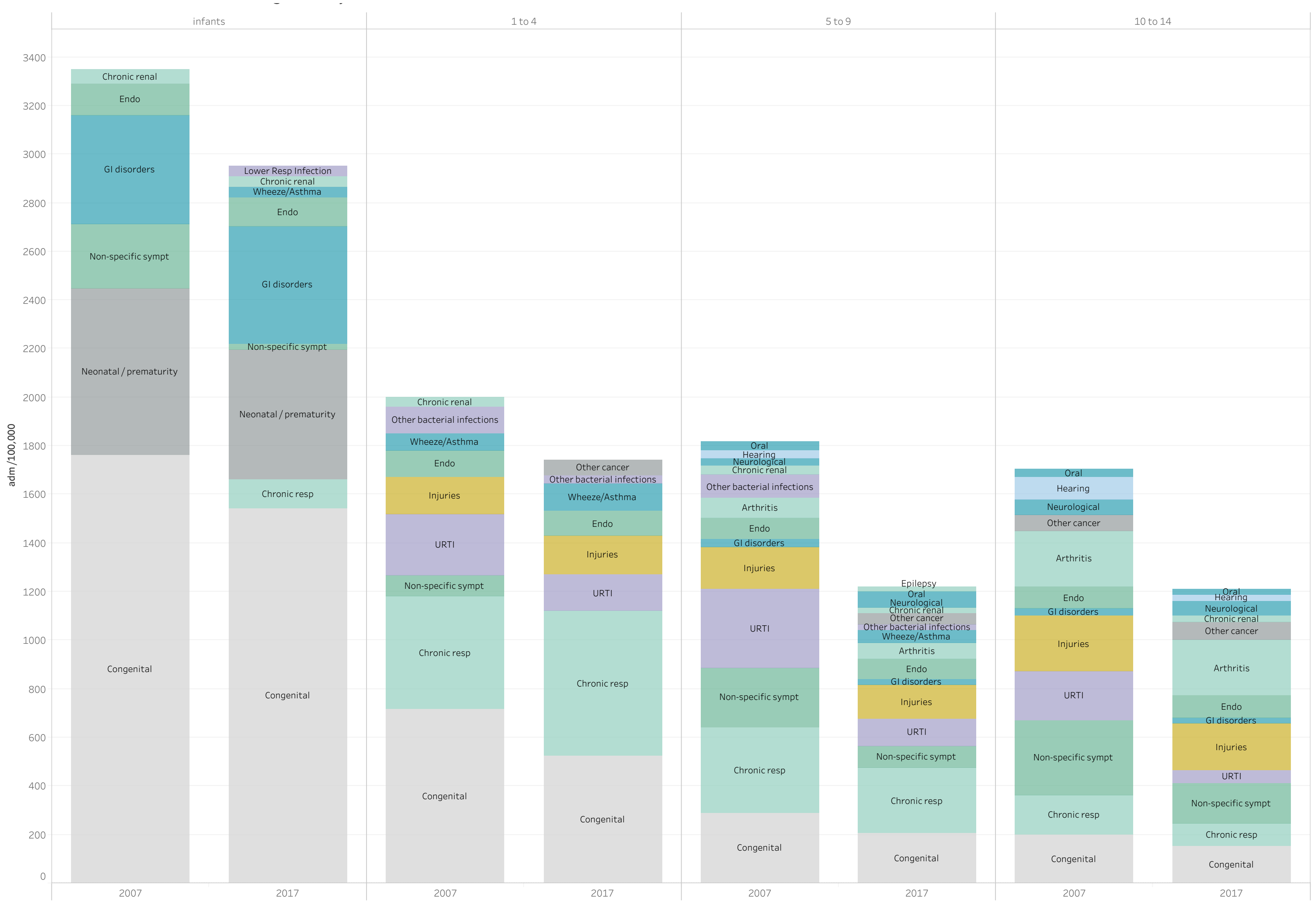

#### Appendix Figure 8. Causes of elective admissions amongst 15-24 yea olds, 2007 and 2017

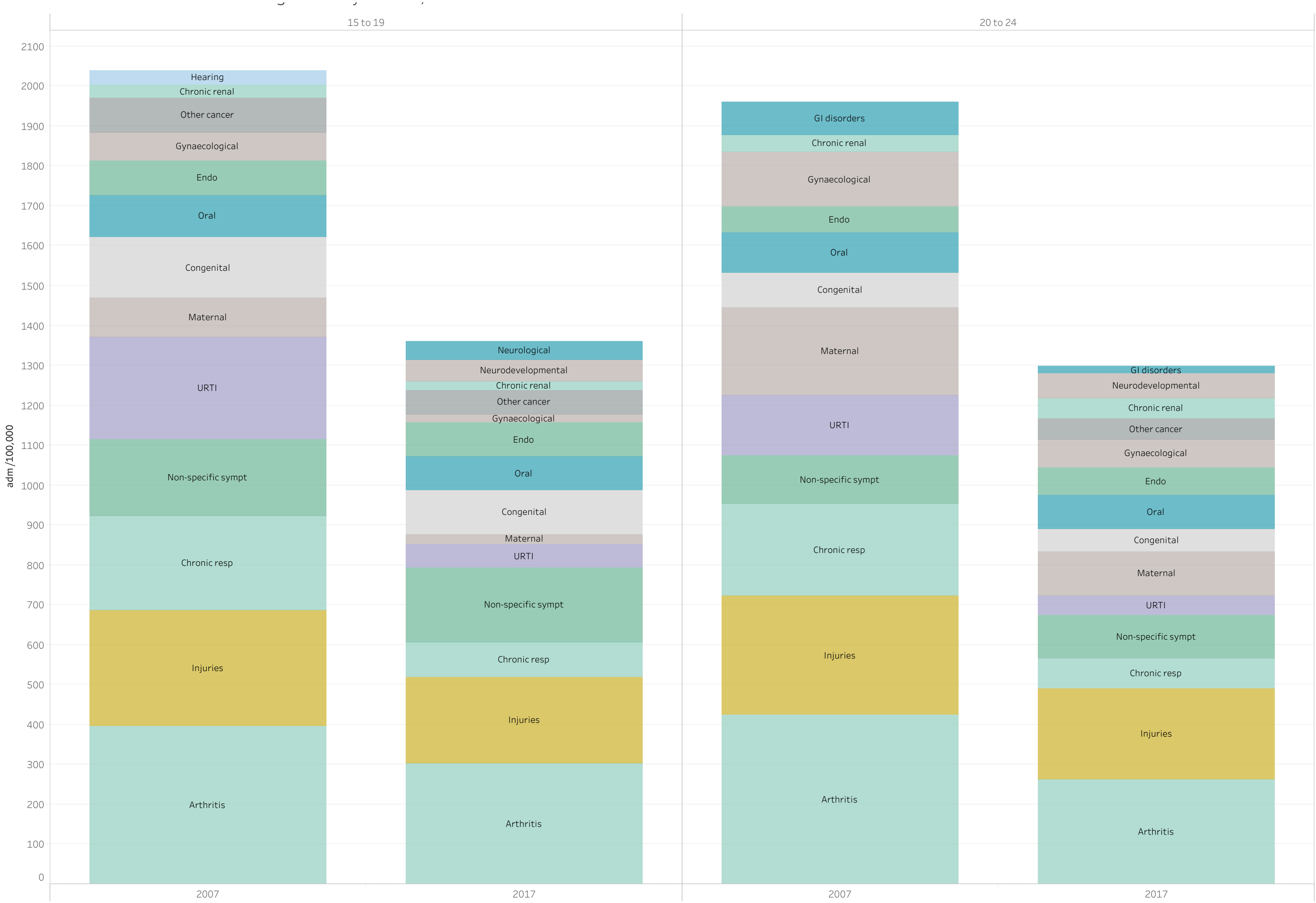

**Appendix Figure 9. Stable poverty scenario 99% CI for model projections for ED attendances and emergency admissions**

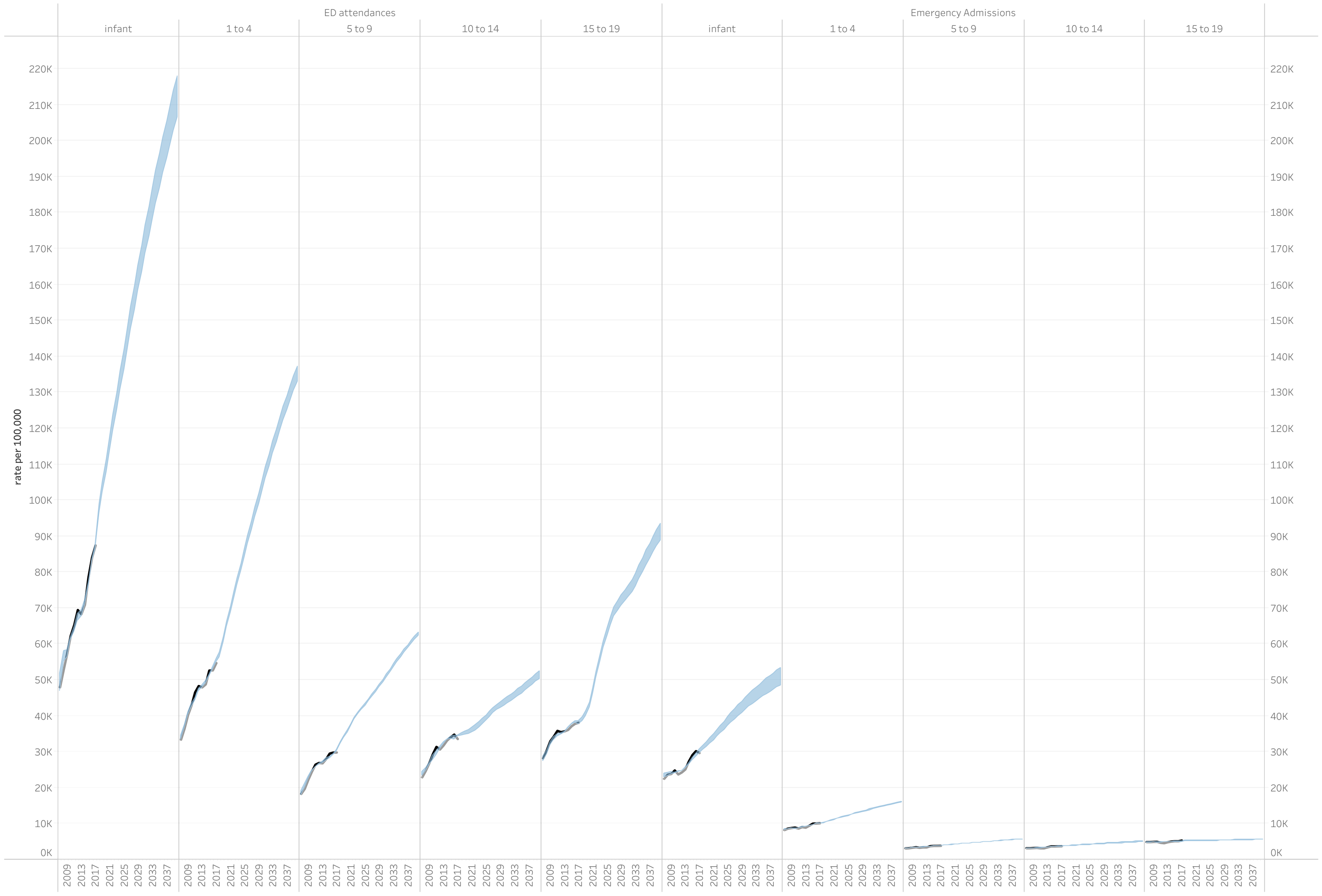

**Appendix Figure 10. Decreasing poverty scenario 99% CI for model projections for ED attendances and emergency admissions**

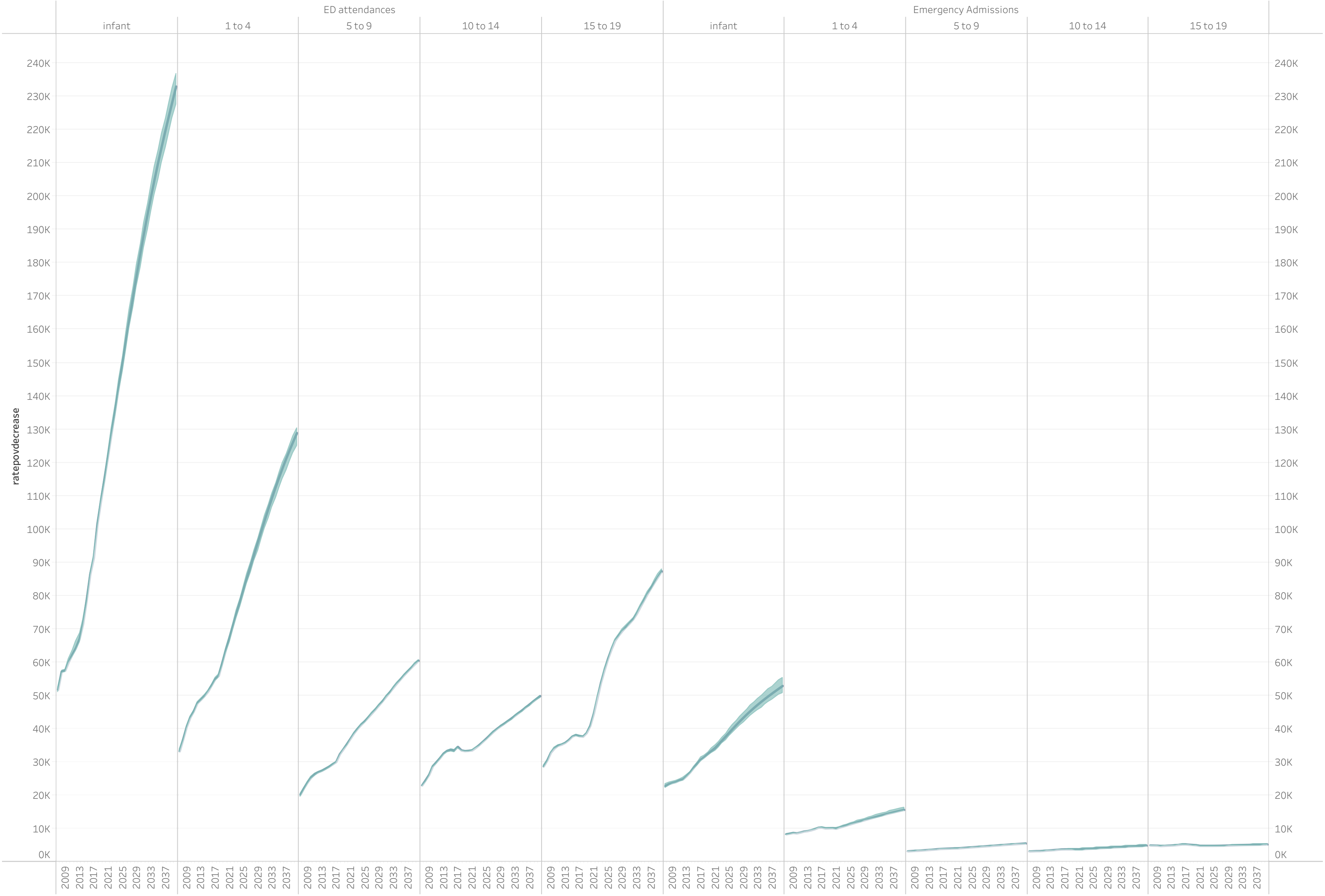

**Appendix Figure 11. Increasing poverty scenario 99% CI for model projections for ED attendances and emergency admissions**

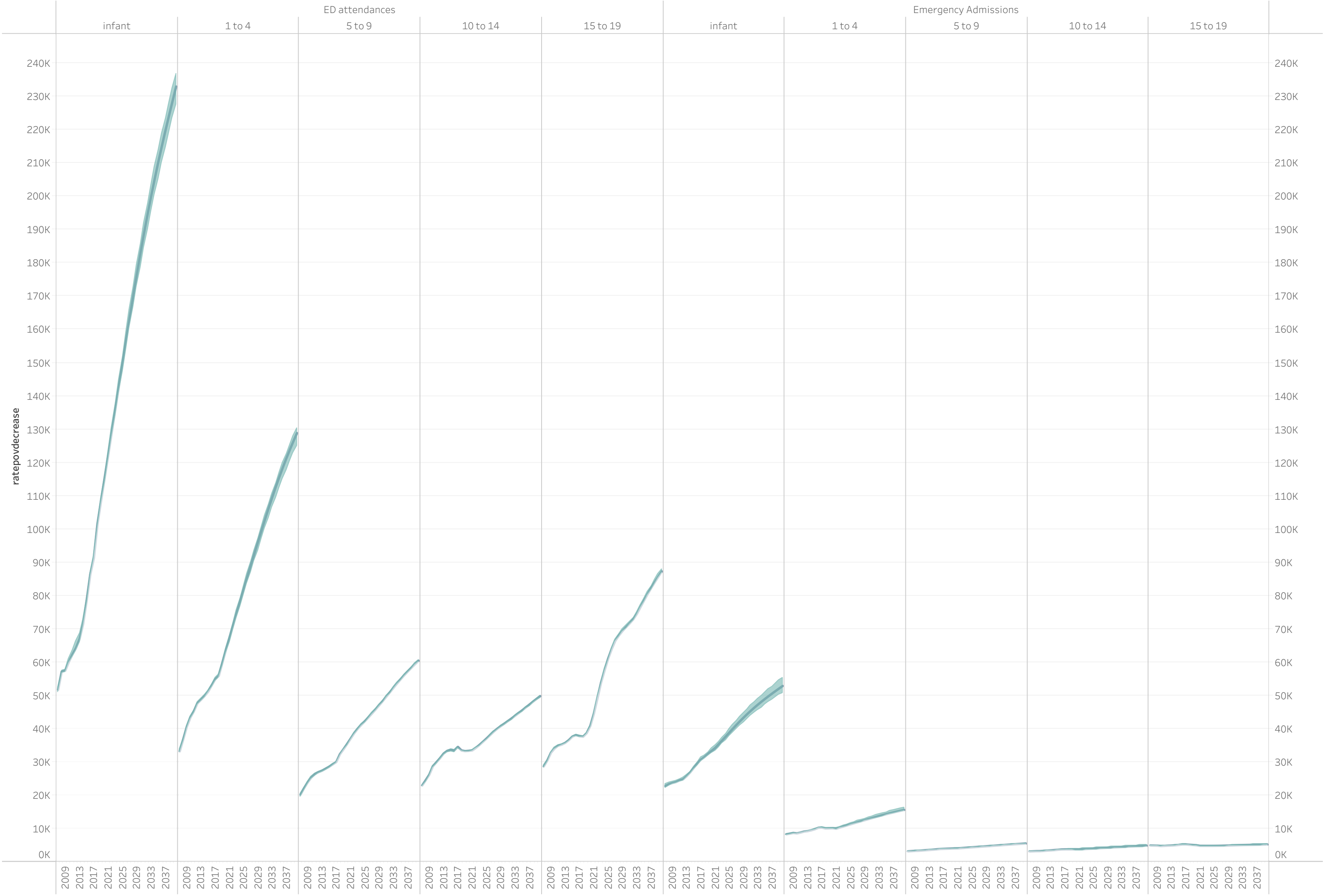

#### Appendix Figure 12. Outpatient attendance rate projections to 2040 by poverty scenario

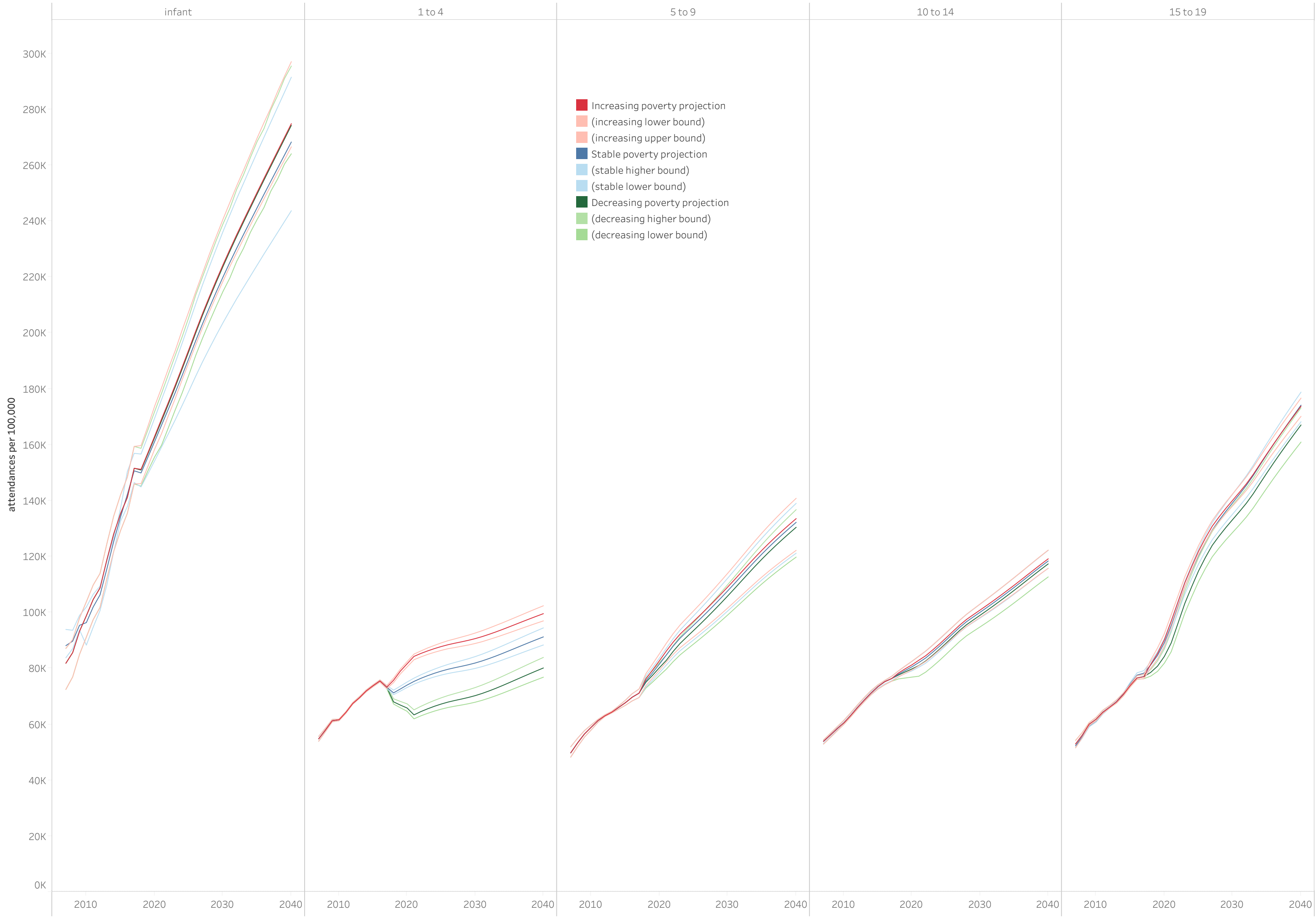
